# A new method for exploring gene-gene and gene-environment interactions in GWAS with tree ensemble methods and SHAP values

**DOI:** 10.1101/2020.05.13.20100149

**Authors:** Pål Vegard Johnsen, Signe Riemer-Sørensen, Andrew Thomas DeWan, Megan E. Cahill, Mette Langaas

## Abstract

**Background:** The identification of gene-gene and gene-environment interactions in genome-wide association studies is challenging due to the unknown nature of the interactions and the overwhelmingly large number of possible combinations. Classical logistic regression models are suitable to look for pre-defined interactions while more complex models, such as tree ensemble models, with the ability to detect any interactions have previously been difficult to interpret. However, with the development of methods for model explainability, it is now possible to interpret tree ensemble models with a strong theoretical ground and efficiently.

**Results:** We propose a tree ensemble- and SHAP-based method for identifying as well as interpreting both gene-gene and gene-environment interactions on large-scale biobank data. A set of independent cross-validation runs are used to implicitly investigate the whole genome. We apply and evaluate the method using data from the UK Biobank with obesity as the phenotype. The results are in line with previous research on obesity as we identify top SNPs previously associated with obesity. We further demonstrate how to interpret and visualize interactions. The analysis suggests that the new method finds interactions between features that logistic regression models have difficulties in detecting.

**Conclusions:** The new method robustly detects interesting interactions, and can be applied to large-scale biobanks with high-dimensional data.

## 1 Introduction

In a traditional genome-wide association study (GWAS) each single nucleotide polymorphism (SNP) is tested individually for association with a particular phenotype. Using computationally efficient generalized linear mixed models that account for population stratification and cryptic relatedness, this approach can successfully identify risk alleles in the genome for complex diseases such as type 2 diabetes, Celiac disease and schizophrenia using large biobanks consisting of hundreds of thousands of individuals and SNPs [26, 50, 53]. Despite this, the estimated effects of the risk alleles are typically small and a large proportion of the estimated genetic heritability is yet to be explained for common traits and diseases [33]. One reason is that most traits and diseases are highly polygenic, and thus many risk alleles with tiny effects will not be declared statistically significant due to stringent *p*-value significance thresholds. A second reason may be failure to account for *epistasis*, namely interactions between genes which together can impact the association with a certain phenotype in a non-linear way [38]. The statistical and biological understanding of epistasis has been discussed to a large extent due to its many possible misinterpretations. In this paper, statistical tests for epistasis will be in accordance with Cordell [12]. A third reason for the missing genetic heritability may be gene-environment interactions where the effect of a variant depends on some external environmental factor.

With the increasing focus on epistasis, many exhaustive search algorithms have been developed such as GBOOST, SHEsisEpi and DSS, and by using graphics processing units (GPUs) [55, 51, 20, 18]. It has been shown that a GWAS investigating pairwise SNP-SNP-interactions with 6 · 10^5^ SNPs and 15,000 samples can be computed in a couple of hours using the aforementioned algorithms [9]. However, it is expected that the number of samples will increase by hundreds of thousands and possibly millions of individuals over the next several years. This will pose serious challenges with respect to memory capacity. In addition, the number of directly genotyped SNPs to evaluate, ignoring imputed genotype values, may be of the order of millions. The number of possible pairwise interactions to investigate will therefore be enormous, while only a small proportion of these combinations may be important with respect to the trait of interest. In addition, inclusion of environmental features is either not considered or limited in the aforementioned exhaustive search algorithms [55]. This can lead to overlooking important relationships. Within modern biobanks, a rich amount of information, clinical, demographic, environmental and genetic, is available for each individual. A GWAS implemented using biobank data should therefore take full advantage of information with any perceived relevance for the trait of interest. However, construction of parametric models incorporating all these features including interactions is difficult. The models will consist of many parameters and assumptions, and the most powerful statistical tests are too computationally intensive in an exhaustive search setting [13]. One way to deal with this difficulty is a two-stage method where the first stage is to find the most influential features, followed by more thorough investigation in the second stage on these features as is done in Li and Won [23] using GBOOST in the first stage. Here, we suggest a similar approach where we first rank the importance of each feature using the tree ensemble model XGBoost [10]. Recent research has demonstrated the possibility to interpret efficiently and with strong theoretical ground the importance of each feature from tree ensemble models using so-called SHapley Additive exPlanation (SHAP) values [32]. Based on this ranking, we further propose a model fitting process where the aim is to find the best XGBoost models with respect to predictive performance. Finally, based on these models, the aim is to explain the relationships that the models consider most important, and specifically the interactions.

By using real data from UK Biobank, we demonstrate these models’ capability to: a) Rank features by importance and thereby removing noise. b) Evaluate the use of XGBoost as both a predictive model and explainable model, and finally c) Rank and explain plausible gene-gene and gene-environment interactions. We finish by comparing the top ranked interactions with classical logistic regression with interaction terms and perform statistical tests. In this paper, the focus is on a case-control setting. Obesity was selected since this particular trait has been extensively researched in previous GWAS [25, 48, 49]. This provides a meaningful way to evaluate our method.

## 2 Background

Recent research within GWAS to account for both genetic and environmental interactions have focused on how to explore the large amount of data in a more systematic way by using various non-parametric machine learning models such as tree ensemble models and deep neural networks [45, 29, 52]. So far, the most successfully applied machine learning methods for genotype data are tree ensemble models such as gradient tree boosting models [43] first introduced by Jerome H. Friedman [14], but with subsequent improvements. One such improvement is the so-called XGBoost implementation [10] used in this paper. XGBoost, as any tree ensemble model, consists of many so-called *weak learners* which in our case are *regression trees*. There are several advantages of using trees as they can naturally handle data of mixed type (continuous, categorical etc.) and missing values, they have the ability to deal with irrelevant and correlated variables, and they are computationally efficient to use [19]. However, trees suffer from low predictive power, high variance, lack of smoothness, and inability to capture linear structures. High variance and overfitting are of greater concern with deeper trees. Tree ensemble models, consisting of many trees, will reduce this variance and improve the predictive power [19]. In fact, smoothness and ability to capture linear structures have also been shown to be improved [35]. In addition, a recent paper published by Lundberg et al. [32] showed that tree ensemble models have the capability to be efficiently and objectively interpreted by measuring the importance of each feature with respect to the predictions of the model by introducing so-called SHAP values. Interpretation of the XGBoost models through SHAP values will allow us to explain the prediction for each individual, a beneficial property in a precision medicine setting.

### 2.1 Problem description and syntax

The aim of a GWAS is to detect associations between a phenotype and one or more single nucleotide polymorphisms (SNPs). Let *y*_*i*_ be the value/phenotype of some trait for individual *i*. This value may signify the absence or presence of a certain trait, such as a disease, or some continuous measure such as height, weight or blood pressure, or even a combination of measures such as the body mass index (BMI). Let *g*_*i,a*_ denote the number of copies (0, 1 or 2) of the minor allele (referred to as the genotype) for a biallelic SNP *a* and individual *i*. Furthermore, let *x*_*i,e*_ denote the value of some environmental feature, and let the matrix **X**_*N×M*_ represent all genetic and environmental data for all N individuals and M features. Usually in a GWAS, the association between a SNP and a trait is tested separately for each SNP using a generalized linear mixed model. However, another approach is to model the association between several SNPs and a trait simultaneously. We will use the latter approach, and will refer to genetic and environmental data as *features*, **x**_*i*_, for each individual *i*. Consider a model for predicting the phenotype, *y*_*i*_, denoted *ŷ*_*i*_(**x**_*i*_). The performance of the model depends on how close each *ŷ*_*i*_(**x**_*i*_) is to *y*_*i*_ for all individuals with respect to some loss function. However, equally important in this setting is to understand what influences the prediction *ŷ*_*i*_(**x**_*i*_). In other words, we would like to understand how *each feature* contributes to the prediction *ŷ*_*i*_(**x**_*i*_) for each individual *i*. The more complex the model is in terms of non-linearity, the more complicated it will be to explain each prediction. However, a non-linear model that can account for several features at once opens the possibility of exploring interactions. In this paper we aim to derive such a model and we will specifically consider the special case where the trait *y*_*i*_ is binary, that is, presence or absence of a phenotype. We denote the group consisting of individuals where the phenotype is absent as the *control group*, and the other group as the *case group*.

### 2.2 XGBoost

XGBoost uses regression trees as building blocks, as illustrated in Figure 1. An important aspect of trees, is that they automatically handle interactions between features. Consider the leftmost tree in Figure 1, where the first split is for feature *x*_1_, and then for both branches of the tree the next split is for feature *x*_2_. Observe that the impact of feature *x*_2_ in the tree is dependent on the value of feature *x*_1_, with a different outcome if *x*_1_ *≤* 1 than if *x*_1_ = 2. This means that a statistical interaction between feature *x*_1_ and *x*_2_ is encoded in the tree.

**Figure 1:**
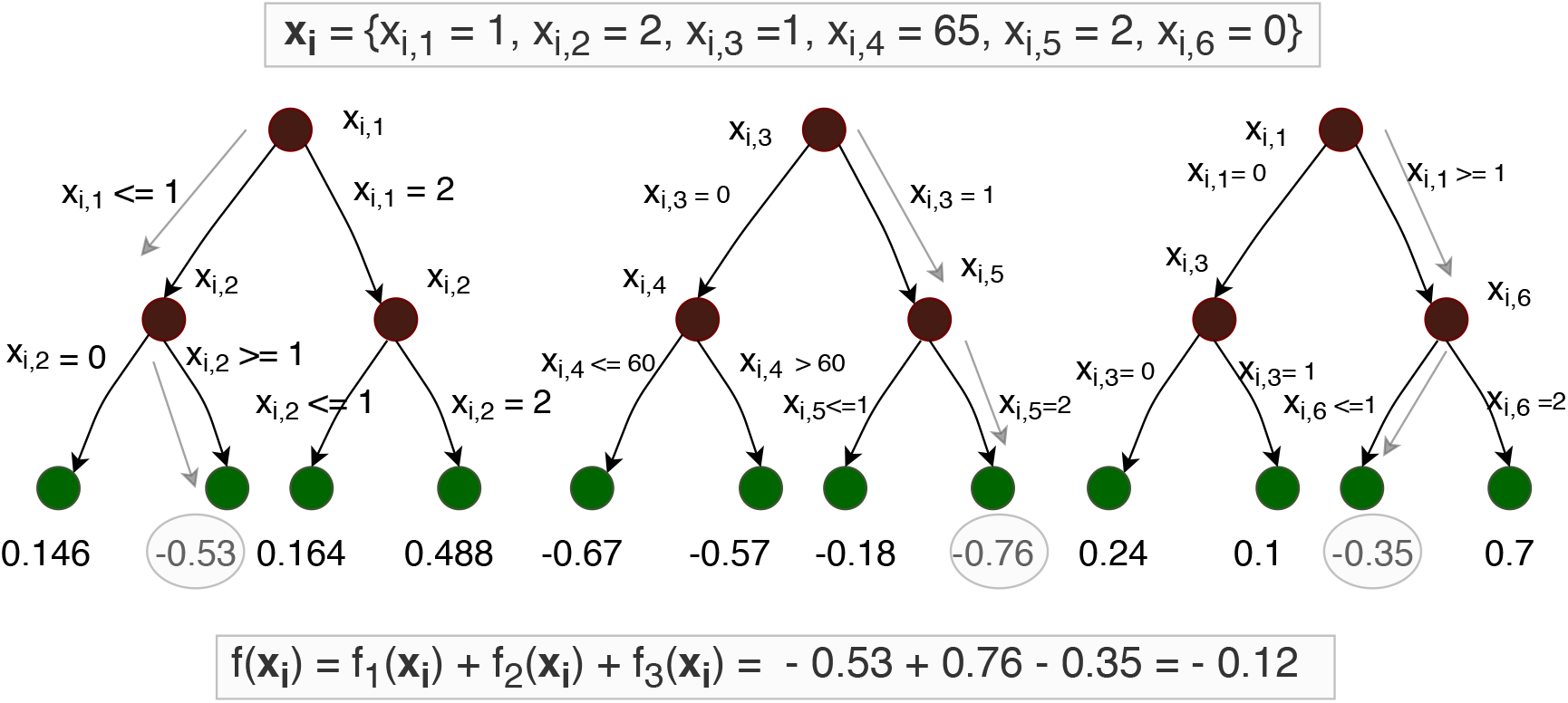
An example with three constructed regression trees with six features *x*_*i*,1_ to *x*_*i*, 6_ used as splitting points at each branch, and leaf node values. Also shown is the computation of f(x_i_) given an example of feature values x_i_. The structure of the trees opens the possibility to explore interactions since a path from a root node to a leaf node denotes a combination of feature values.

#### 2.2.1 Constructing trees

The XGBoost algorithm starts with the construction of a single regression tree, and then new regression trees are consecutively constructed. The construction of each tree is based on information from the former trees. After building *t −* 1 trees for the training data matrix **X**_*N×M*_, the total loss function *L*^(*t−*1)^(**X**_*N×M*_) is given by:

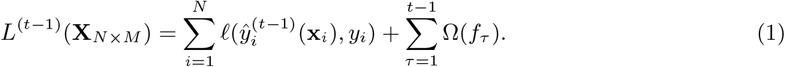

The loss function per individual,. *ℓ*(), is a differentiable convex function which measures the performance of the prediction, 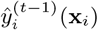, with respect to the observed trait, *y*_*i*_, for an individual *i* with features **x**_*i*_ when there is a total of *t −* 1 trees in the model. In a binary classification setting (e.g. case versus control), a convenient loss function is the binary cross-entropy:

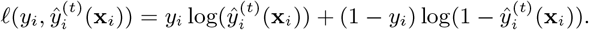

Regression tree number *τ* is denoted as *f*_*τ*_, a data structure that contains information of nodes, features used as splitting points and leaf node values. The function *f*_*τ*_ (**x**_*i*_) *∈* ℝ in (1) outputs the value of the leaf node (green circles in Figure 1) corresponding to features **x**_*i*_ based on tree *τ*. In a binary classification setting, the prediction 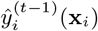 is interpreted as the probability that individual *i* is a case given *t –* 1 regression trees.

In order for 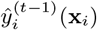 to represent a probability given the *t−*1 regression trees, a much used transformation is the sigmoid function:

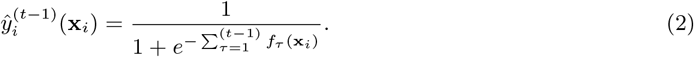

The function Ω in (1) is a regularization factor that penalizes the loss function by the number of nodes, *T*_*τ*_, in tree *τ* and leaf node values *w*_*τk*_, *k ∈ {*1, ‥, *T*_*τ*_ *}*:

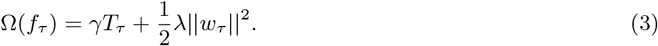

As is the standard procedure in any gradient tree boosting method, the next regression tree 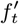 is constructed to minimize the total loss function:

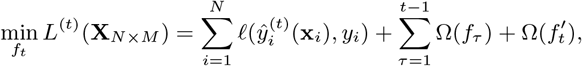

where

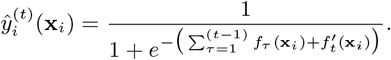

Using second order Taylor approximation of the loss function. *ℓ*(), it can be shown that for a given tree structure, the values of the leaf nodes that minimize the total loss function can be computed [10]. When constructing each tree, one starts at the root node and successively investigates which feature to use as a splitting point at each node. The model will choose the split that minimize the total loss function at that point. There are different strategies when constructing the trees. Either one can make a split at the leaf node, out of several, that minimize the total loss, or one can split at the leaf node that is closest to the root node. Splitting at the node which gives the largest decrease in loss is the approach that will be used in our case. Even finding the optimal split greedily is time-consuming and memory-inefficient in the case of hundreds of thousands of features. The XGBoost R software package applies the histogram method to reduce the search time [2, 22, 24]. For the handling of missing values, we refer the original XGBoost paper [10].

The model will typically stop training when the total loss function has not decreased in a given number of iterations, where a new regression tree is constructed in each iteration. The prediction of the final model on the logit scale given features **x**_*i*_ and a total of *T* trees is given by 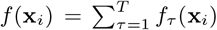, while the probability of the case class will be calculated using the sigmoid transform on *f* (**x**_*i*_), as in Equation (2).

#### 2.2.2 Hyperparameters in XGBoost

XGBoost has a large set of hyperparameters, which may influence the performance of the algorithm and its ability to find the best representation of the data. In this paper, we focus on the learning rate *η, subsample, colsample bytree, colsample bylevel* and *max depth*. The learning rate *η ∈*(0, 1] scales the values of the leaf nodes after the construction of each new tree, in which case 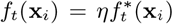 where 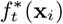 is the raw regression tree before the scaling of the leaf node values has been applied. This will limit particular trees to dominate the prediction. It has been shown to be important since it governs how fast the model will learn and it can prevent early overfitting. In high-dimensional problems this is crucial and the learning rate should be well below 1 and is typically 0.1 or smaller [5, 19]. The subsample and colsample hyperparameters decide the proportion of individuals and features to be evaluated in each regression tree. They also prevent overfitting, and in addition reduce the training time of the model. A typical value for both hyperparameters is 0.5, and in high-dimensional data it has been proposed that even smaller values can be beneficial [19]. However, this will depend on what proportion of the high-dimensional data is relevant. If the relevant proportion is small, a more reasonable value is closer to 1 [10]. The parameter colsample bytree is used to partition the number of possible features to use as splitting points in each level of the tree. The literature is quite scarce on its effect, but it may oppose the non-optimal greedy approach search as well as providing more room for learning in a way similar to the learning rate. Other important hyperparameters are the regularization parameter *λ* already seen in Equation (3), as well as the parameter early stopping rounds which is the maximum number of rounds without predictive improvement of the validation data before the training stops. To avoid overfitting, the validation data is independent of the training data.

### 2.3 Classification performance metric

For a binary classification model, the predictive performance in the validation data can be evaluated with specific focus on the group that is of particular interest (the case group). Let TP, FP and FN be the number of true positives (true cases classified as cases), false positives (controls falsely classified as cases) and false negatives (cases falsely classified as controls), respectively. The precision and recall given the predictions from a model are defined as follows,

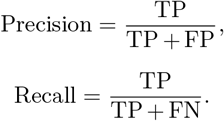

The model prediction, *ŷ*_*i*_ *∈* (0, 1), for each *i* is converted into a classification rule by choosing a cut-off, often set to 0.5, and for the given cut-off the prediction is either a TP, FP or FN. Instead of using a single cut-off, the strength of the prediction model can be evaluated by computing precision and recall for different cut-offs, and plotting the precision-recall pairs to give a precision-recall-curve. The better the model is, the closer the pair of precision and recall measures will be to the point (1,1) for each cut-off, and so a measure for the model performance can be the area under the curve, denoted PR-AUC (precision-recall area-under-curve). PR-AUC is most often used in the case of imbalance, meaning that one group is larger than the other. When TP = 0 and FP = 0, corresponding to a model that always predicts an individual to be a control, the precision is defined to be zero.

### 2.4 A measure of feature importance in tree ensemble models - SHapley Additive exPlanation (SHAP) values

When evaluating the global feature importance in a tree ensemble model, one possibility is to look at the relative decrease in loss for all splits by a given feature over all trees [6]. Unfortunately, this measure suffers from so-called *inconsistency* as pointed out in Lundberg, Erion, and Lee [30]. In short, this means that the feature contributions are unfairly distributed as a result of not accounting for the importance of the order in which the features are introduced in the trees. Another popular, but similarly inconsistent, importance metric is counting the number of times each feature is used as a split point. Instead, a metric based on so-called SHapley Additive exPlanation (SHAP) values can be shown to achieve consistency [31, 32]. In this case, each feature *j* for each individual *i* is given a SHAP value, *ϕ*_*i,j*_ (**x**_*i*_), which represents the contribution of feature *j* with respect to the prediction, 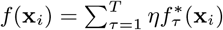, equal to the output of the linear sum of all *T* regression trees in a tree ensemble model given features **x**_*i*_. This metric exhibits several favourable properties aside from consistency [31]. For instance, the sum of the contributions of each feature, *ϕ*_*i,j*_ (**x**_*i*_), including a constant *ϕ*_0_ equals the prediction of the model *f* (**x**_*i*_):

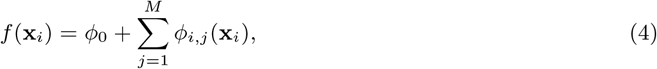

where *M* is the number of features included in the model. Moreover, the total contribution of a subset of all features for each individual is simply equal to the sum of the SHAP values for each feature. The reason for these favourable properties is that the contribution, *ϕ*_*i,j*_ (**x**_*i*_), is computed based on a concept from game theory first introduced by Lloyd Shapley [44]:

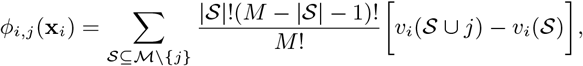

where ℳ is the set of all features included in the model, the function *v*_*i*_(𝒮) measures the total contribution of a given set of features (*v*_*i*_(ℳ) = *f* (**x**_*i*_)), and the sum is across all possible subsets where feature *j* is not included. The parameter *ϕ*_0_ is defined as *ϕ*_0_ = *v*(𝒮 = ∅). The key idea is that the contribution of each feature for each individual is measured by evaluating the difference between the prediction when the value of feature *j* is known, versus the case when the value feature *j* is unknown for all subsets 𝒮 *⊆* ℳ *\ {j}*. In a statistical setting, the *marginal expectation* first introduced in Janzing, Minorics, and Blöbaum [21] is shown to be a reasonable measure of *v*_*i*_(𝒮):

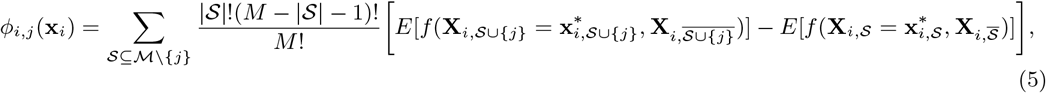

where 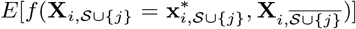 is the expected prediction when only the values of the feature subset 𝒮 as well as feature *j*, denoted 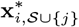, are known, while the vector of the complement set, 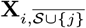, is regarded as a random vector. Notice that 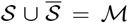. The values *ϕ*_*i,j*_ (**x**_*i*_) are denoted as SHAP (SHapley Additive exPlanation) values [31]. In the case of binary classification using a tree ensemble model, the prediction *f* (**x**_*i*_) can be interpreted as the log-odds prediction.

By assuming all features are mutually independent, Lundberg et al. [32] constructed an algorithm to estimate the SHAP values given in Equation (5) in polynomial running time, *O*(*T LD*^2^), with maximum depth *D* and maximum number of leaves *L* in all *T* trees. The assumption about mutual independence is a limitation, and without this assumption the estimation of the SHAP values becomes more complicated [1]. For further details about estimations of SHAP values assuming mutual independence, see Supplementary File.

#### 2.4.1 SHAP interaction value

The SHAP values can be further generalized to interpret pairwise interactions through the SHAP interaction values Φ_*i,j,k*_(**x**_*i*_), *j ≠ k*, for individual *i* and features *j* and *k* given by [15, 32]:

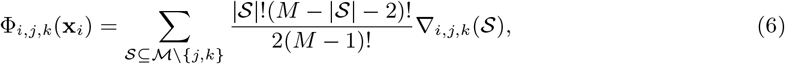

where

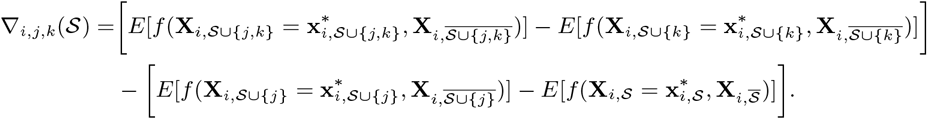

If feature *k* yields additional information when present simultaneously with feature *j, ∇*_*i,j,k*_(𝒮) will be different from zero with the sign depending on how feature *k* (when present) affects feature *j*. With these definitions, the pairwise SHAP interaction values have the same properties as the single-feature SHAP values. For instance, the contribution of a given feature *j, ϕ*_*i,j*_ (**x**_*i*_), can be separated into the contribution of *j* itself, denoted Φ_*i,j,j*_(**x**_*i*_), in addition to all interactions including feature *j*, denoted as Φ_*i,j,k*_(**x**_*i*_), for all *k ≠ j*:

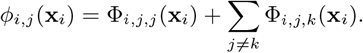

The final prediction for each individual can be decomposed into

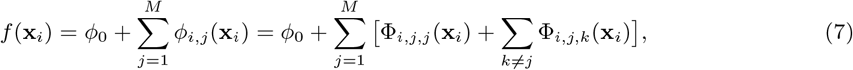

where Φ_*i,j,k*_(**x**_*i*_) = Φ_*i,k,j*_(**x**_*i*_).

The interactions for all possible pairs of features for a particular tree ensemble model can be computed in *O*(*T MLD*^2^) time [32].

## 3 Tree ensemble- and SHAP-based method for identifying interactions

We propose a new method using XGBoost and SHAP values to identify interactions between features, that is, either SNP-SNP interactions or SNP-environment interactions. The method is outlined in Figure 2.

**Figure 2:**
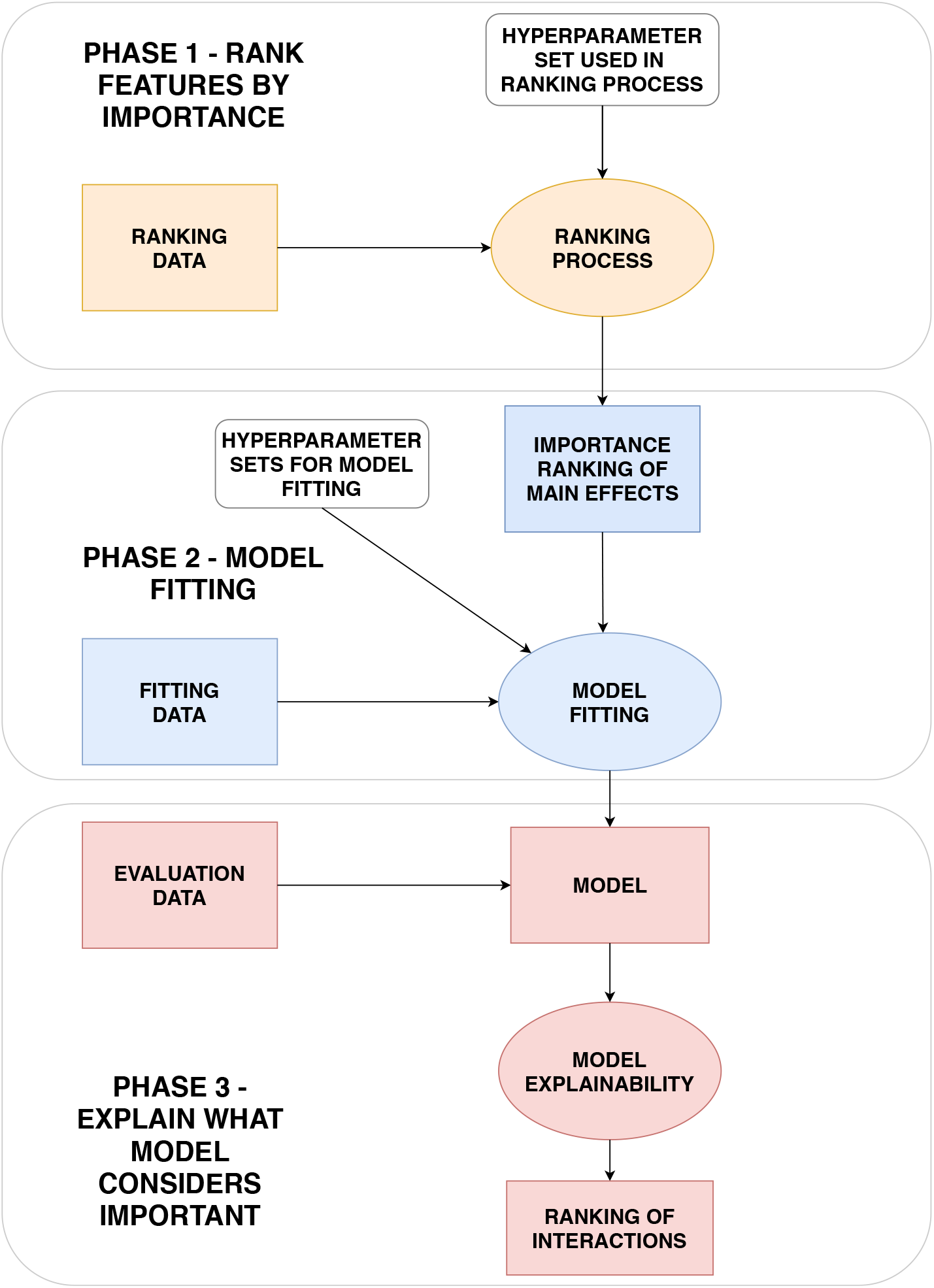
The ranking, model fitting and explanation phases. In the ranking phase, the SNPs and environmental features are ordered by their relative importance. The ranking is achieved with XGBoost and SHAP values as explained in Section 3.1 and Figure 4. In the model fitting phase, the top ranked features are combined and modelled with XGBoost as described in Section 3.2 and Figure 6. Finally, the explanations and interactions are obtained from the SHAP values. This is discussed in Section 3.3 and visualized in Figures 10, and 11.

**Figure 3:**
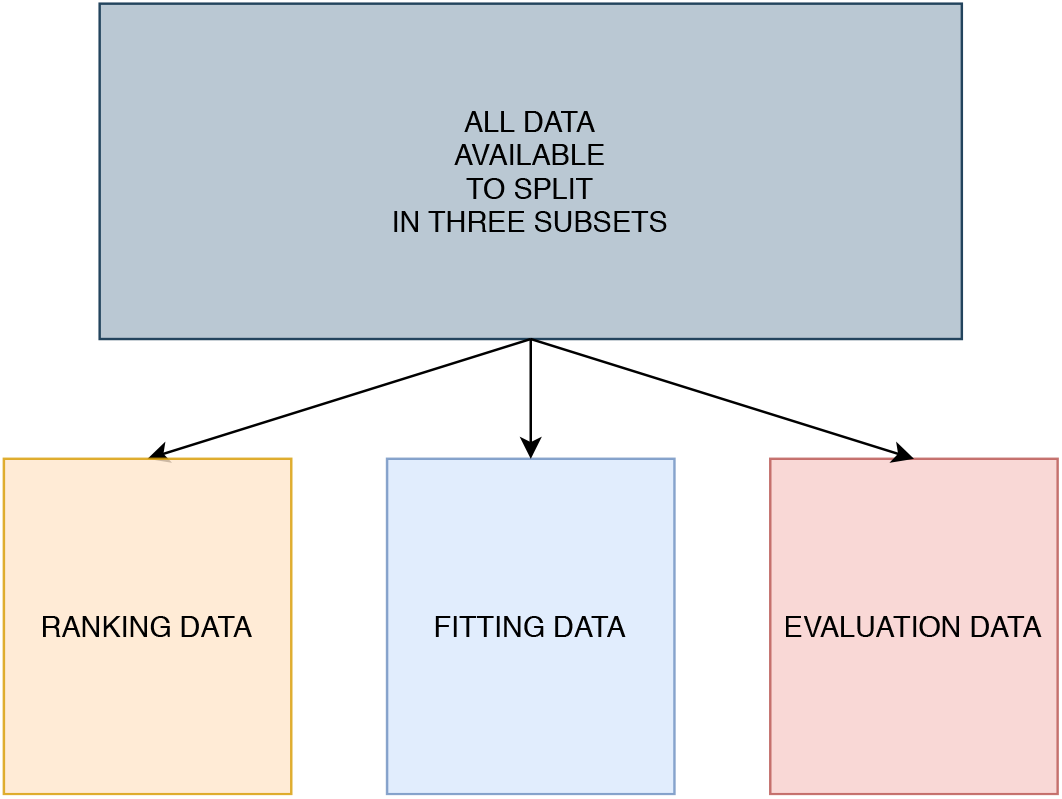
All data available is divided into three subsets: Ranking data, fitting data and evaluation data. The ranking data is used to rank features by importance in order to remove noise. The fitting data is used to fit models by using the ranking derived from the ranking data. The evaluation data is finally used to explain what is considered important with respect to the predictions from the models trained on the fitting data.

We use a tree ensemble model, such as XGBoost, trained on data consisting of observations from individuals each with a phenotype *y*_*i*_ and features **x**_*i*_, to rank features by importance using SHAP values. The ranked list of features makes it possible to construct new models that use only the most important features, and therefore have higher predictive power. Finally, having a fitted model that only consists of relevant features, we want to explain which relationships are important with respect to the phenotype, both marginal effects and interactions.

In order to evaluate the ability to both rank features by importance, find the best predictive models, and explain the best models without causing optimism bias, we divide the individuals in three disjoint subsets, namely the *ranking data, fitting data* and *evaluation data*.

Dividing the data into several subsets will reduce the power to detect relevant features as well as reducing the degree to which each subset is representative of the full dataset. However, the procedures are intended to be used on data from large biobanks to reduce power loss and representativeness of the subsets. By using independent subsets of the data for each phase of our method, we avoid potential overfitting by reusing data, and will be able to give an accurate account to which extent tree ensemble models are able to capture relationships between features and the trait of interest that classical GWAS methods might have difficulties to achieve [3].

### 3.1 Phase 1: The ranking process

Identifying associations between SNPs and a phenotype is a typical example of a high-dimensional problem. Experience from several GWAS suggests that many low-effect SNPs are not detected. At the same time we still expect only a small proportion of the total genome to have any effect with respect to the trait of interest. Consequently, we face a challenge where many potential SNPs have a causal effect on the trait, but a much larger number of SNPs are not causal at all and therefore contribute as noise. To make it even more complicated, among the large number of SNPs in the human genome, there exist correlations between different SNPs throughout the whole genome in a given population called *linkage disequilibrium* [42]. In general, the closer the physical distance between a pair of SNPs is, the more correlated the SNPs tend to be. As not all SNPs are genotyped, and if we disregard imputed data, there will be gaps between the SNPs that are present. We expect that in many cases, SNPs with causal effect fall in such gaps. But here we are helped by the linkage disequilibrium and the correlation between nearby SNPs. For practical purpose this means that a subset of all SNPs available can provide information beyond only those SNPs selected, but also those nearby SNPs that are in linkage disequilibrium. This also applies for interactions.

The analysis is further complicated by confounders such as population stratification and cryptic relatedness between individuals which can lead to spurious associations in our models [47]. cross-validation is a model validation technique in which several models of identical structure are trained on different portions of the training data, and each model is evaluated on independent validation data. With respect to feature importance, a procedure with the purpose of preventing spurious associations, is to evaluate the importance of each feature based on all models constructed during cross-validation.

From our knowledge about linkage disequilibrium, population stratification and cryptic relatedness, we therefore propose a method to implicitly investigate the whole genome efficiently and objectively through a series of independent cross-validations by using XGBoost, a tree ensemble model, as shown in Figure 4. It is from these independent cross-validations we will provide a ranking of the importance of each feature.

**Figure 4:**
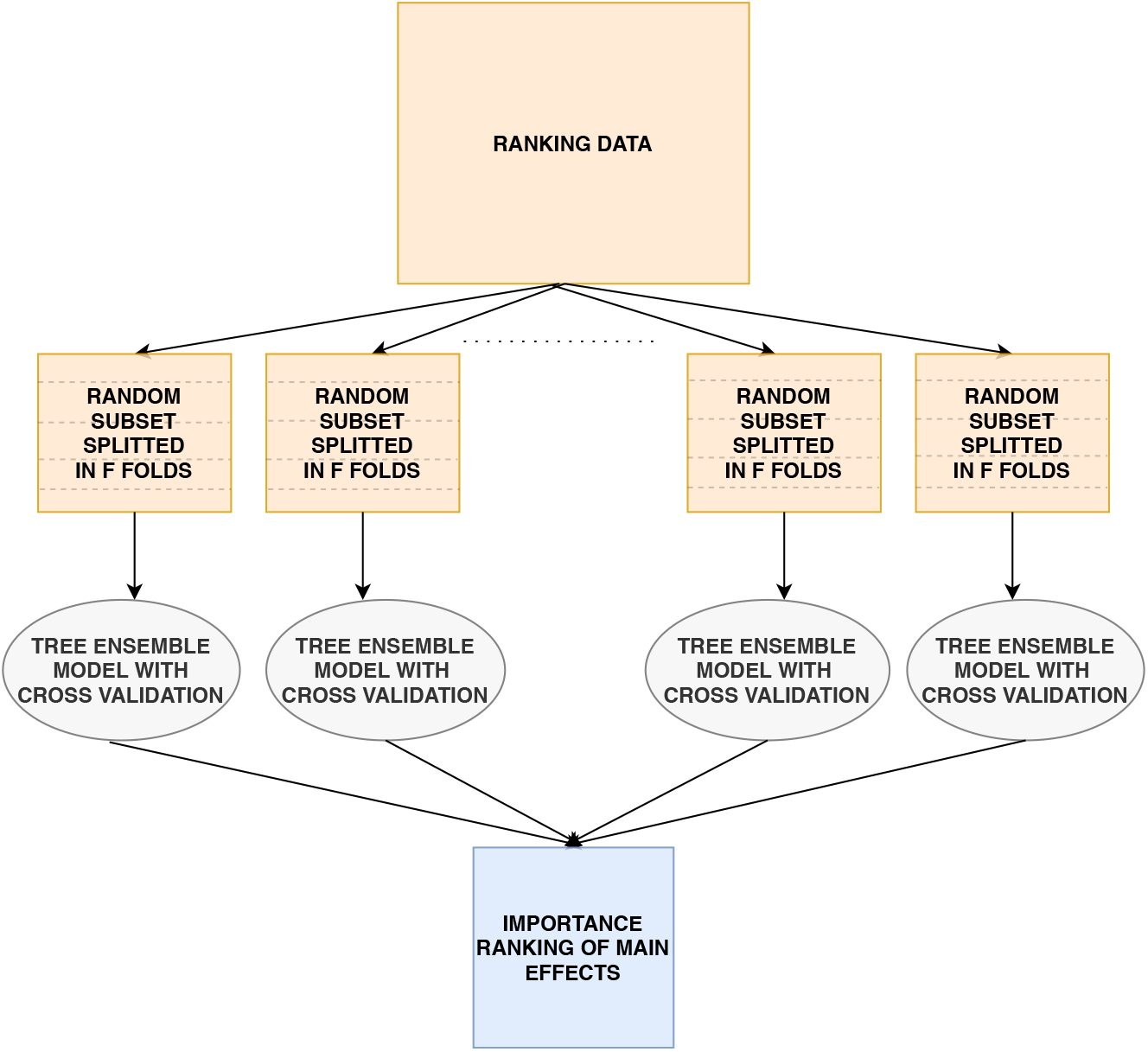
In the ranking process, multiple independent subsets are created and used in a cross-validation procedure with tree ensemble models. The trained models will be used to rank the importance of the features.

Consider a dataset with *N* individuals and *R* directly genotyped SNPs. We create *A* randomly selected subsets, where each subset consist of *S* SNPs with low mutual correlation and *G ≤ N* individuals randomly sampled with equal probability in order to keep an as agnostic narrowed search as possible. Furthermore, each sampled subset is divided into *F* folds where *F −* 1 folds are used in an ordinary cross-validation to train *F −* 1 XGBoost models, while the last fold never seen or used during cross-validation is used as test data. This will create *F −* 1 models trained on different data, and their performance can be objectievly evaluated on the test data. As shown in Figure 5 for the *F −* 1 folds used in cross-validation, in each iteration *F −* 2 folds are used to train an XGBoost model, while the last fold is used as validation data. Training of the model will proceed as long as the performance on the validation data improves within a certain number of iterations as given by the early stopping rounds hyperparameter discussed in Section 2.2.2. Cross-validation reduces the harm of both overfitting and selection bias [34]. The degree of overfitting can be further investigated by looking at the model performance difference on the validation and test data.

**Figure 5:**
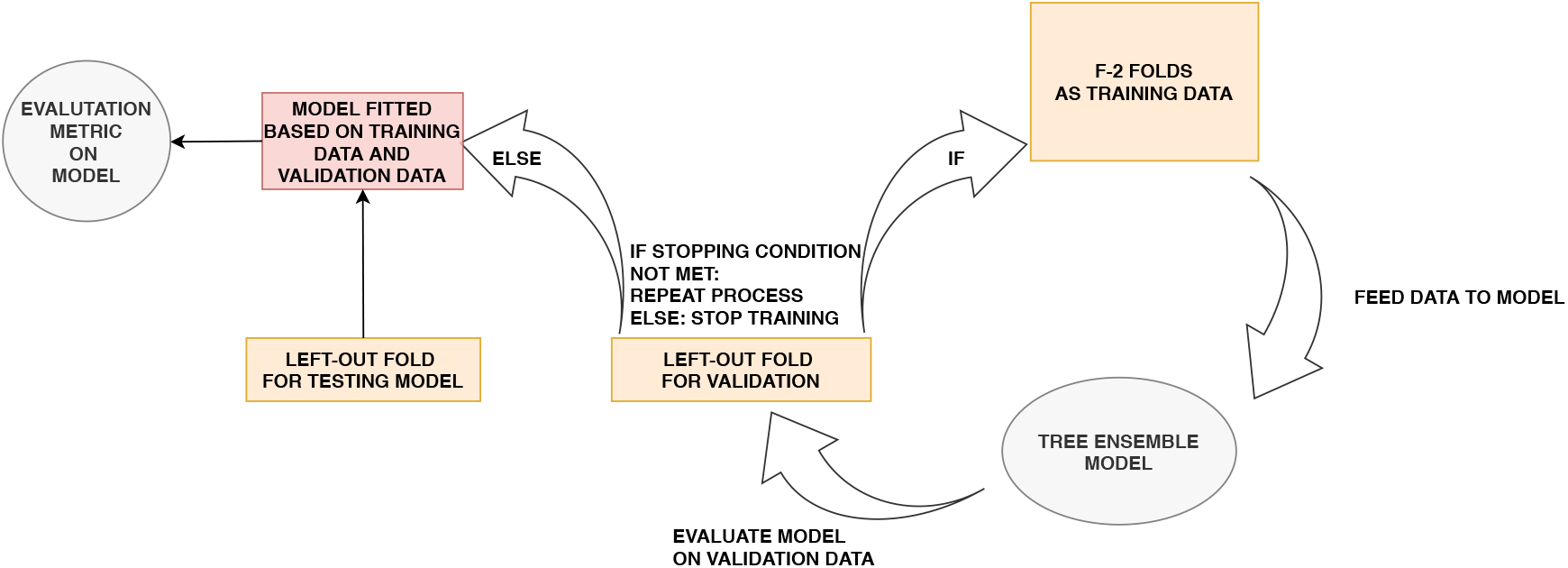
The cross-validation phase when training data consists of *F −* 2 specific merged folds. Training of the model will proceed as long as the performance on the validation data improves within a certain number of iterations as given by the early_stopping_rounds hyperparameter.

With *A* subsets each creating *F −* 1 models, the question is now how to rank all features investigated in all *A* subsets for all *𝒫* = *A*(*F −* 1) models. We define the *relative feature contribution*, denoted 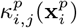, for individual *i*, feature *j* and model *p* as:

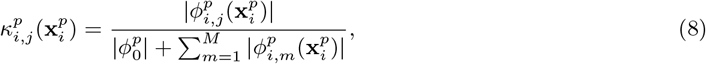

where 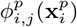 is the SHAP value for feature *j* from model *p* with feature values 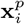. The measure 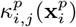can be interpreted as the proportion of the prediction for individual *i* attributed to feature *j* for model *p*. We now want to estimate the expected relative contribution of feature *j* using all the past independent cross-validations. The expected relative feature contribution (ERFC), *Ê*[*κ*_*j*_], is estimated as:

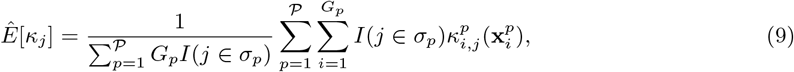

where 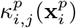 denotes the relative feature contribution of feature *j* for individual *i* in a set of *G*_*p*_ individuals used to explain model *p*, and *I*(*j ∈ σ*_*p*_) is the indicator function which is equal to one if feature *j* is included in the subset data used to train model *p*, and zero elsewhere.

The individuals *G*_*p*_ used to explain a particular model *p* created from a particular subset *a* are chosen to be the individuals from the test data of the subset. This means that the contribution of each feature in each model will be based on individuals never seen during training. The estimation of *Ê*[*κ*_*j*_] for each feature *j* will finally create a ranking of the contribution of each feature.

### 3.2 Phase 2: The model fitting process

Given a ranked list of features based on their feature contribution with respect to some trait, this allows for avoidance of irrelevant features and increases the ability to detect important relationships.

At this stage we are interested in finding the models with the best performance on some test data by utilizing the ranking of feature importance from the ranking process. For this purpose we use data never seen before in order to avoid any optimism bias, by using the fitting data [3]. The heterogeneity as well as possible relatedness among the individuals are taken into account by again using cross-validation. First we split the data in *F* folds, of which *F −* 1 folds are used for cross-validation while the last fold is used as test data. This gives *F −* 1 fitted models in total. The model fitting procedure is summarized in Figure 6 which shows how one model, out of *F −* 1, is fitted using only the top *K* features as well a set of hyperparameters. The aim is to find which set of *F −* 1 models that on average performs best on the test data as a function of the value of *K* and hyperparameter values.

**Figure 6:**
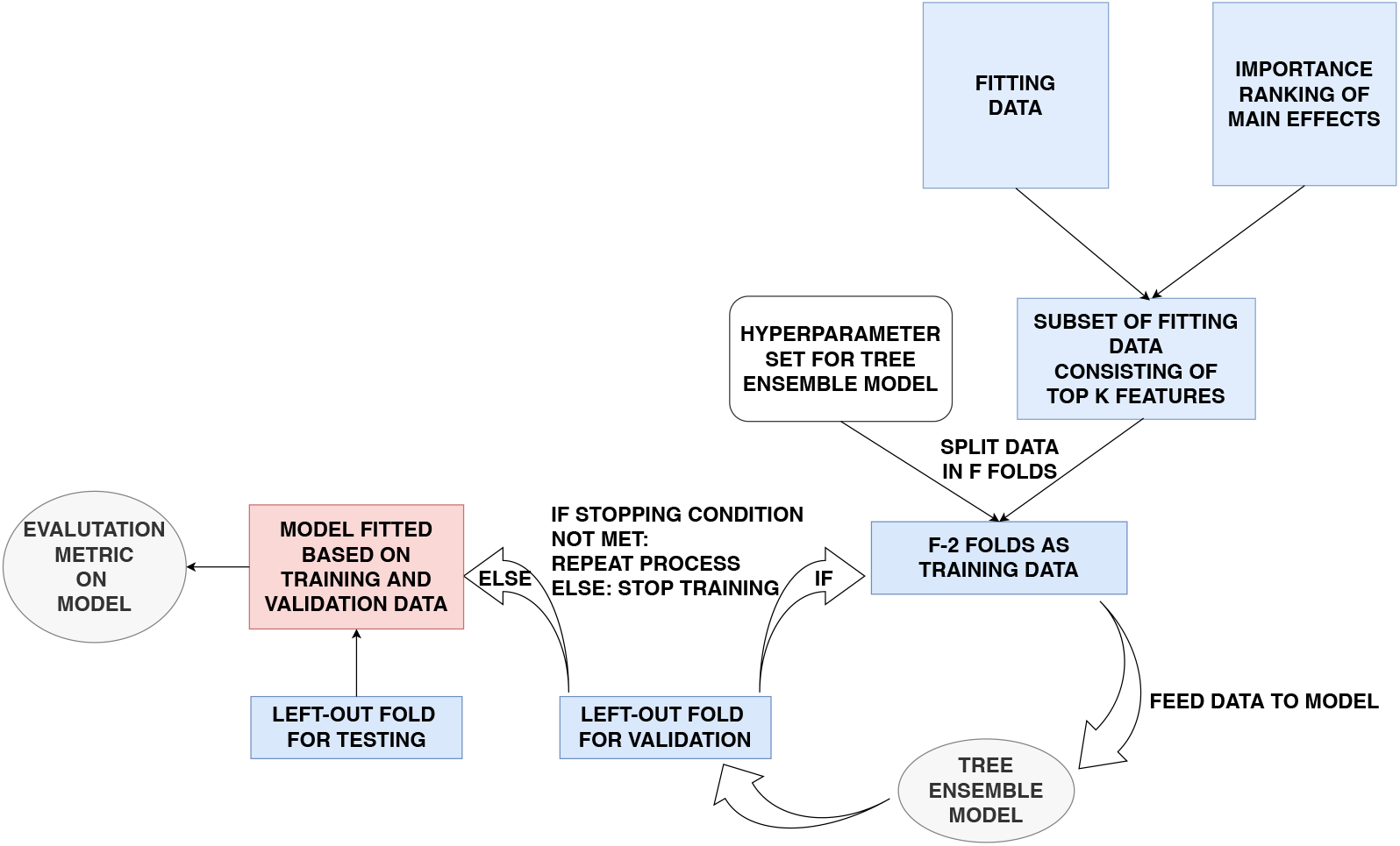
Given a table of ranked feature importances, XGBoost models based on the top *K* features are trained in a new cross-validation procedure based on an independent set of individuals, namely the fitting data. We search for the XGBoost models that on average performs the best for a given set of hyperparameters (including the value of *K*) based on test data.

In order to explain the XGBoost models at a later stage we want to compute the SHAP values. We assume the features are mutually independent when computing the SHAP values. To take this into account, we combine the ranking with low values of the mutual squared Pearson’s correlation, denoted *r*^2^, when selecting the *K* features to include. See Section 2 in Supplementary File for more information. Even though we are not guaranteed an independent set of features using *r*^2^, it significantly limits the number of dependent features and therefore reduces the negative effect of misleading computations of SHAP values.

### 3.3 Phase 3: Model explainability

After finding the best predictive models from the model fitting process, we can investigate which features and interactions contribute to the models through the SHAP values. Along the same lines as for the marginal feature importance in Section 3.1, the *relative contribution* for each interaction between feature *j* and *k* for a particular individual *i* and model *p* can be computed as:

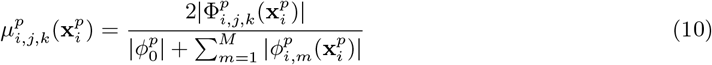

We can estimate the expected relative interaction contribution, 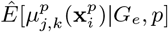, given data consisting of *G*_*e*_ individuals and a model *p*:

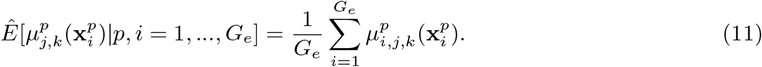

The *G*_*e*_ individuals are part of the evaluation data as shown in Figure 2. As we have *F −* 1 models from the model fitting process, we average the result from all *F −* 1 models:

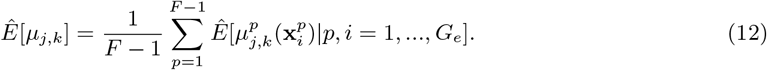

We define this as the expected relative interaction contribution (ERIC). This will provide a ranked list of interactions. A ranked list of marginal effects can be achieved in the same way as described in Section 3.1, but this time based on the *F −* 1 models constructed after the model fitting process.

The contribution of the top ranked marginal effects and interactions to the prediction for each individual can be visualized with sina plots and partial dependence plots as illustrated in Figure 10 and 11 [32]. For one particular trained tree ensemble model, the sina plot in Figure 10 shows the SHAP value for each individual indicated as a point with color depending on the value of the feature. The larger the absolute SHAP value, the more the feature contributes to the model prediction for a specific individual. Partial dependence plots, exemplified in Figure 11, are used to visualize how the contribution, in other words the SHAP value, for a particular feature depends on another feature for different combinations of feature values. Here as well, each individual is marked as a point with the value of a given feature given on the x-axis and the corresponding SHAP value for this feature with respect to the prediction on the y-axis. The color of the point, however, represents the value of some other feature. In this way, interactions can be visualized and interpreted.

## 4 Example: Application using UK Biobank data

As an example, we apply and evaluate the method described in Section 3 to real data from the UK Biobank Resource [7]. Among the available phenotypes, obesity was examined because it has been subjected to a number of high quality and well-powered GWAS that have identified more than 100 loci, many that have been consistently replicated across studies (e.g. FTO, BDNF, MC4R, TMEM18, SEC16B) [25, 48, 49]. Thus, we have a good set of true-positive loci with which to compare our results. We only analyzed White European individuals to limit the effect of population stratification. The method outlined in this work can be applied to both continuous and discrete phenotypes, but as explained in Section 1, in this example we consider a binary trait. We define an individual to be part of either the control group (*y*_*i*_ = 0) or case group (*y*_*i*_ = 1) by:

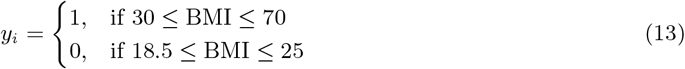

As should be evident above, we exclude overweight individuals with 25 *<* BMI *<* 30 from the analysis and only compare normal-weight individuals (18.5 *≤* BMI *≤* 25) with obese individuals (BMI *≥* 30). This reduces the number of subjects available for analysis, but allows us to define more distinct case and control groups. For power analyses of extreme phenotype data we refer the reader to [4]. The BMI data is provided from measurements at the initial assessment visit (2006-2010) at which participants were recruited and consent given. Phenotype-independent quality control of the genetic data for White European subjects consisting of the genotyped SNPs is completed using PLINK1.9 [41], and the details are given in Appendix A. We only consider directly genotyped SNPs. In addition, we limit our analysis to SNPs with minor allele frequency (MAF) greater than 1%. By only considering the two groups defined in Equation (13), this results in a total of 529 024 SNPs and 207 015 individuals to investigate, of which 43% of these individuals are in the group defined as obese.

### 4.1 Environmental covariates

We will include environmental features that are preivously reported to be of importance with respect to obesity, namely sex, age, physical activity, intake of saturated fat, sleep duration, stress and alcohol consumption [37, 39, 28, 46, 8]. These environmental features are a representative set for the demonstration of the methodology and were not intended to be an exhaustive set of environmental covariates available in the UK Biobank for obesity. Information about the environmental features, including their definitions, are included in Appendix B.

### 4.2 Ranking, fitting and evaluation data

Following the data split in Section 3, the ranking data consist of 80 000 randomly chosen individuals, and will be used to rank the features by importance. The fitting data also consist of 80 000 individuals. This subset is used to find the best predictive models in the model fitting process. The evaluation data consists of 47 015 individuals, and is used to explain what the models found in the model fitting process consider the most important features and in which way they contribute. In all subsets, we retain the proportion of obese individuals.

### 4.3 Phase 1: The ranking process

By using the ranking data, at this stage we create *A* = 50 subsets where each subset consists of *G* = 70000 individuals and *S* = 110000 randomly chosen SNPs corresponding to 21% of the total number of SNPs available. The choice of total number of subsets to create is motivated from Equation (2) in Supplementary File with the criteria that any pair of SNPs appears in the same subset at least once with 90% certainty. The larger the number of individuals in each subset, the higher statistical power, but at the same time, the memory capacity limits the number of individuals in each subset at the cost of lost power. As the ranking process is time-consuming, we do not attempt any sophisticated hyperparameter optimization, but instead choose four hyperparameters sets that we regard as reasonable, given in Table 1. In addition, in all further analysis, the regularization parameter *λ* is set to 1, the default value in most XGBoost softwares [11]. The parameter early stopping rounds is set to 20.

**Table 1:**
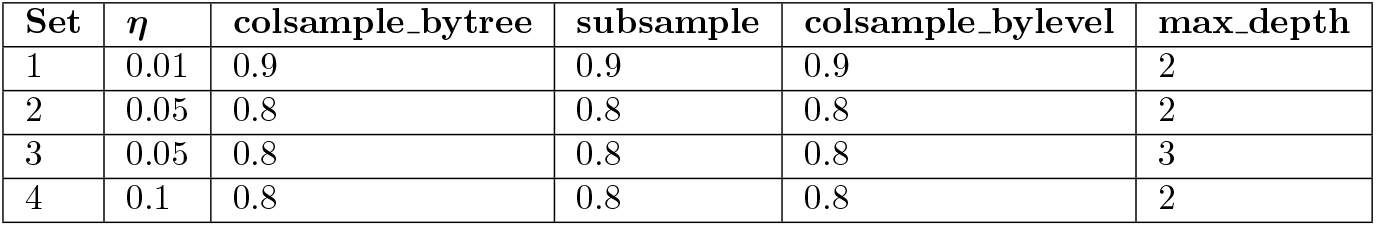
The four hyperparameter sets for XGBoost considered in the analysis during the ranking process described in Section 3.1.

As discussed in Blagus and Lusa [5], the learning rate *η* is set to be small for high-dimensional data such as 0.1, while as discussed in Chen and Guestrin [10], colsample bytree is set to be large as there is only a small proportion of all features that are relevant. The hyperparameter subsample is also set to be large in order to increase the power to detect features of importance. The parameter colsample bylevel has not been extensively discussed in the literature, but the parameter will oppose the greedy construction of the trees which may be beneficial in the long run. The maximum depth of the trees are set to no more than three, the reason being both computational considerations as well as the fact that the marginal expectations used to compute the SHAP values in (5) will be more inaccurate the deeper the trees are (see Supplementary File).

We apply the R package xgboost to both train xgboost models and to estimate SHAP values [11].

Using Equation (9) to estimate the expected relative contribution for each feature as described in Section 3.1, we give the ranking for the top 20 features in Table 2 for hyperparameter set 2 in Table 1.

**Table 2:** The resulting ranking based on the expected relative feature contribution (ERFC) from the ranking process for hyperparameter set 2 in Table 1. The environmental features are, as expected, considered more important than the SNPs, while the most important SNPs are at or nearby the FTO gene in agreement with previous studies.

| Feature | ERFC |
| --- | --- |
| Sex | 0.12 |
| Alcohol intake frequency | 0.12 |
| Physical activity | 0.11 |
| Saturated fat intake | 0.058 |
| Stressful events | 0.056 |
| Sleep duration | 0.049 |
| Age at initial assessment | 0.047 |
| rs17817449 (FTO, Chr. 16) | 0.025 |
| rs1421085 (FTO, Chr. 16) | 0.025 |
| rs1121980 (FTO, Chr. 16) | 0.024 |
| rs7202116 (FTO) | 0.023 |
| rs9941349 (FTO) | 0.023 |
| rs9940128 (FTO) | 0.023 |
| rs9922619 (FTO) | 0.023 |
| rs13393304 (FAM150B - TMEM18, Chr. 2) | 0.022 |
| rs12149832 (FTO) | 0.021 |
| rs9939609 (FTO) | 0.021 |
| rs9930506 (FTO) | 0.021 |
| rs11642841 (FTO) | 0.020 |
| rs2947411 (Chr. 2) | 0.019 |

Not surprisingly, the environmental features are considered most important. The next features are predominantly those connected to the FTO gene at chromosome 16 as expected from previous studies. A SNP close to the TMEM18 gene (rs13393304) is also found in the top 20 list. The next SNPs on the list are predominantly from chromosome 2, one SNP from chromosome 1 at the SEC16B gene (rs10913469) and further down SNPs from chromosome 18, yet no SNPs connected to the MC4R gene for instance. By further investigation, this is due to the fact that the SNPs randomly selected from the 50 subsets did not include any SNPs close to the MC4R gene which illuminates the issue when not creating enough subsets. Apart from this, one can see that the ranking process is able to detect small effects, and importance of each feature can be evaluated by computing SHAP values.

We compare with the corresponding ranked list derived using BOLT-LMM, a Bayesian mixed model that evaluates the marginal effect of each SNP, and computes *p*-values based on the BOLT-LMM infinitesimal mixed-model statistic [26]. The *p*-values computed have been shown to be valid as long as the MAF of each SNP is larger than 1%, and that the case fraction is larger than 30% for a sample of 50 000 individuals [27]. All these criteria are satisfied in our ranking dataset (with case fraction 42 %, MAF greater than 1 % and 80 000 individuals). Table 3 shows the top ranked 13 SNPs (top environmental features are not listed) where features with the smallest *p*-values are regarded to be of most importance.

**Table 3:** The result after running BOLT-LMM on the ranking data showing the top SNPs with smallest *p*-value from the BOLT-LMM infinitesimal mixed-model statistic. All top SNPs are connected to the FTO gene.

| Feature | BOLT-LMM $p$-value |
| --- | --- |
| rs1421085 (FTO) | 3.7E-57 |
| rs9940128 (FTO) | 1.8E-54 |
| rs1121980 (FTO) | 2.4E-54 |
| rs3751812 (FTO) | 7.0E-54 |
| rs17817449 (FTO) | 8.5E-54 |
| rs9939609 (FTO) | 1.3E-53 |
| rs8050136 (FTO) | 2.2E-53 |
| rs7202116 (FTO) | 5.7E-53 |
| rs9941349 (FTO) | 5.0E-52 |
| rs12149832 (FTO) | 3.0E-50 |
| rs9922619 (FTO) | 1.0E-48 |
| rs9930506 (FTO) | 1.1E-48 |
| rs11642841 (FTO) | 1.3E-40 |

In this case, all SNPs are related to the FTO gene, and most of the SNPs except two are also present in Table 2. These two SNPs were not sampled in any subset from the ranking process. The ordering in Table 2 and 3 between SNPs related to the FTO gene are slightly different. However, at this stage it is not strictly necessary to find the true order of the feature impacts, but an approximate order that allows us to discard features with insignificant impact in the further analysis.

### 4.4 Evaluation of the trained models used in the ranking process

To explore the degree of overfitting of the models trained during the ranking process, the PR-AUC score of each model computed on its corresponding validation data and test data (see Figure 5) are explored in a Bland-Altman (mean—difference) plot. This shows the average PR-AUC score for each model on the x-axis, and the difference between the two scores on the y-axis. The results for all chosen sets of hyperparameters given in Table 1 can be seen in Figure 7.

**Figure 7:**
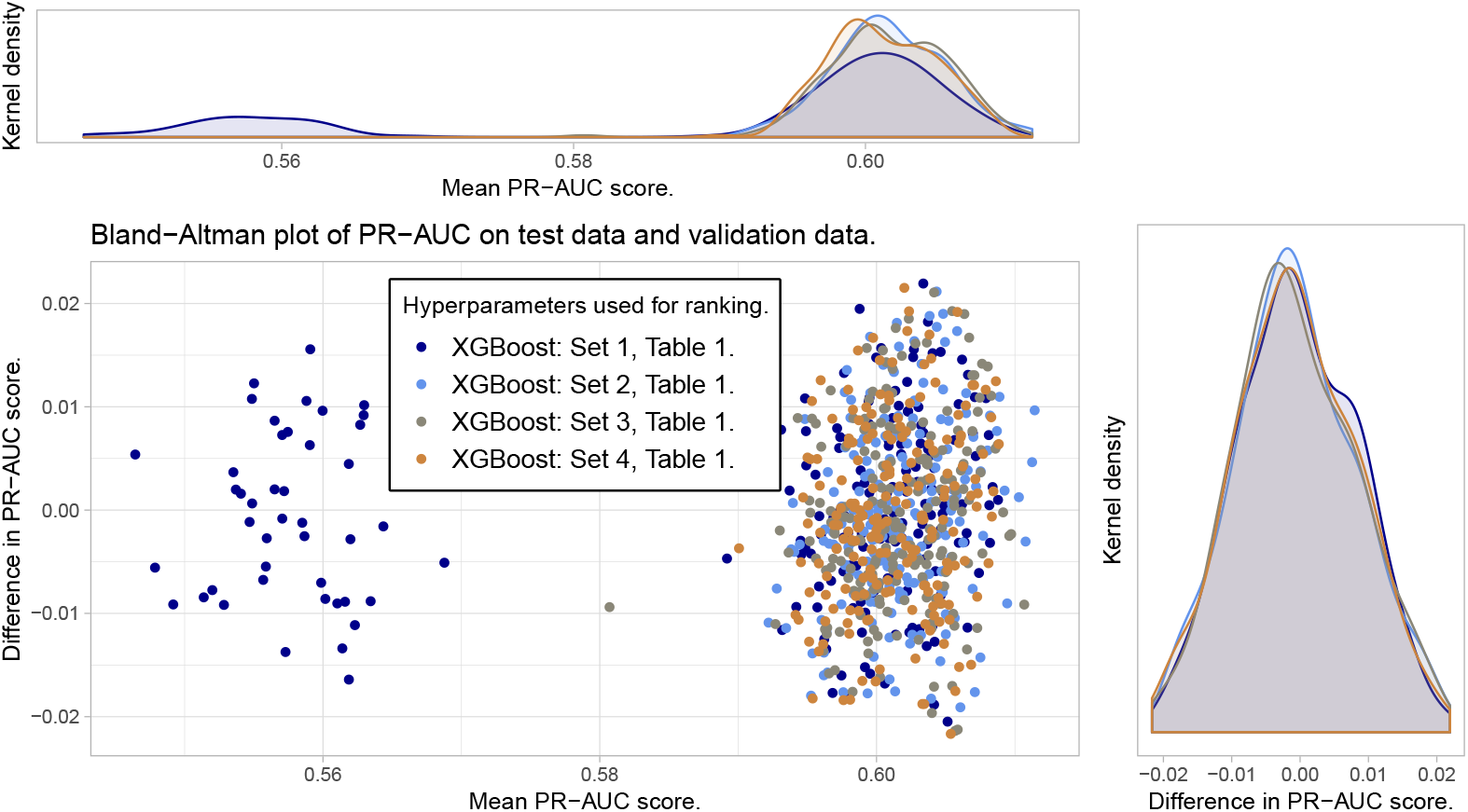
Bland-Altman plot for the trained models used for ranking. No clear signs of overfitting, but one set of hyperparameters shows one cluster of poorer predictions than the others.

Figure 7 shows no clear pattern of overfitting as can be seen from the agreement between the density plots of the difference in PR-AUC scores. However, hyperparameter set 1 from Table 1 shows a cluster of bad predictions with PR-AUC around 0.56. The reason for this can be seen in Figure 8 where bad predictions using hyperparameter set 1 is due to early stopping in the training. When there is no early stopping in the training, we also see that due to the small learning rate given in set 1, more trees are constructed than for the other hyperparameter sets, but yet the performance score is not superior. This emphasizes the importance of hyperparameters.

**Figure 8:**
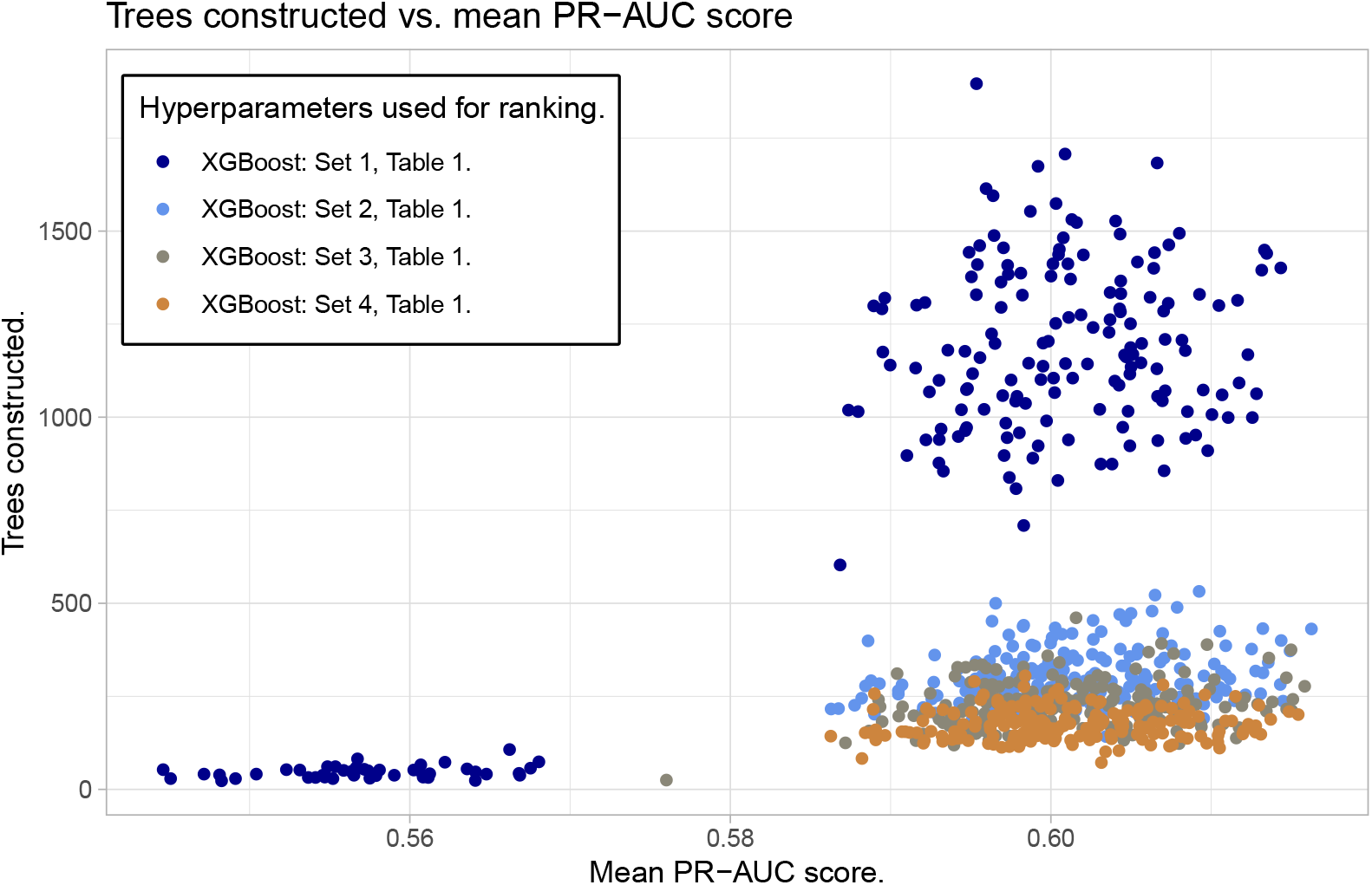
The reason some models with hyperparameter set 1 in the ranking process underperforms is early stopping of the training. Notice also that a larger number of trees need to be constructed to get the same performance as for models with other hyperparameter sets.

### 4.5 Phase 2: Model fit from the ranking process and from BOLT-LMM ranking

As described in Section 3.2, we use the fitting data to train new XGBoost models with cross-validation by including the *K* most important SNPs for *K* = 0 (only including environmental features), *K* = 100, 500, 1000, 3000, 5000, 10000 and finally *K* = 15000. The ranking of the features is the output of the ranking process. In addition, to assess the quality of our method, we also train models based on the ranked table produced by BOLT-LMM as presented in Section 4.3.

Before training, the set of the *K* chosen SNPs is reduced such that the SNPs have mutually squared Pearson’s correlation *r*^2^ *<* 0.2 (see Supplementary File for practical details about implementation). Due to computational limitations, we will only consider hyperparameter tuning from the XGBoost models through the sets given in Table 4, and optimize based on these sets. For each *K* and for the ranking based on our method and the ranking based on the BOLT-LMM model, the maximum average PR-AUC-score for the XGBoost models constructed in the cross-validation is found among the possible hyperparameter sets. For each *K*, we compare how the predictive model perform on the held-out test data from the fitting data. The results are shown in Figure 9. When we vary *K* from small to large values, we expect that the model performance increases the most at the beginning as the most influential features are included, while as more features with low importance are added, the performance increases steadily until it flattens. At the end, the performance may even decrease as noise are added to the model in the form of SNPs without any predictive power.

**Table 4:** The hyperparameter sets considered during Phase 2: Model fitting process, described in Section 3.2.

| Set | $\eta$ | colsample_bytree | subsample | colsample_bylevel | max_depth |
| --- | --- | --- | --- | --- | --- |
| 1 | 0.1 | 0.3 | 0.3 | 0.3 | 2 |
| 2 | 0.1 | 0.5 | 0.5 | 0.5 | 2 |
| 3 | 0.1 | 0.5 | 0.5 | 1 | 2 |
| 4 | 0.1 | 0.8 | 0.8 | 0.8 | 2 |
| 5 | 0.1 | 1 | 1 | 1 | 2 |
| 6 | 0.05 | 0.5 | 0.5 | 0.5 | 2 |
| 7 | 0.05 | 0.8 | 0.8 | 0.8 | 2 |
| 8 | 0.2 | 0.5 | 0.5 | 0.5 | 2 |
| 9 | 0.1 | 0.3 | 0.3 | 0.3 | 3 |
| 10 | 0.1 | 0.5 | 0.5 | 0.5 | 3 |
| 11 | 0.1 | 0.5 | 0.5 | 1 | 3 |
| 12 | 0.1 | 0.8 | 0.8 | 0.8 | 3 |
| 13 | 0.1 | 1 | 1 | 1 | 3 |
| 14 | 0.05 | 0.5 | 0.5 | 0.5 | 3 |
| 15 | 0.05 | 0.8 | 0.8 | 0.8 | 3 |
| 16 | 0.2 | 0.5 | 0.5 | 0.5 | 3 |

**Figure 9:**
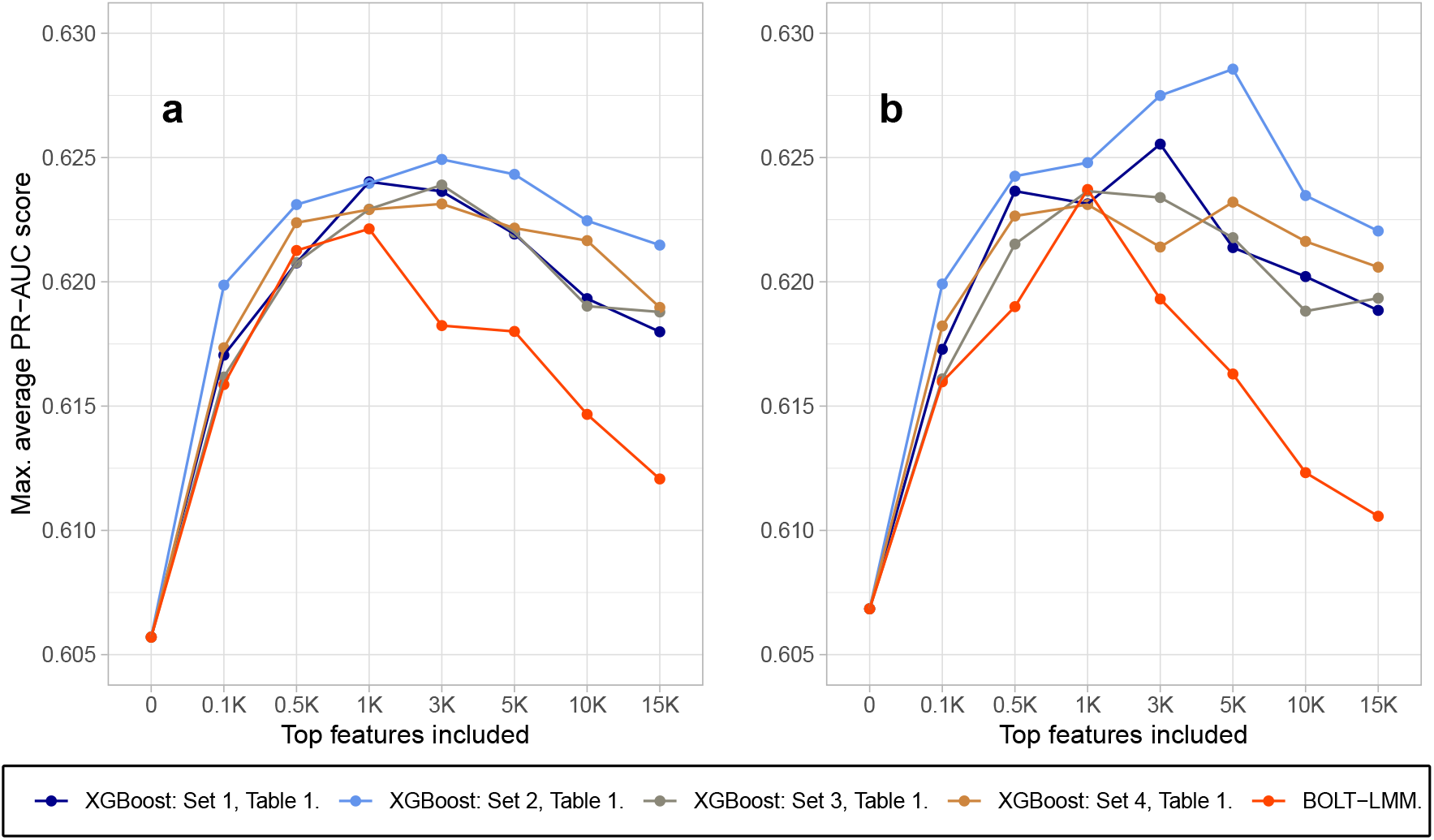
The model fitting process based on top *K* features from both the ranking process (for different sets of XGBoost-hyperparameters indicated by the different colours and the legend) and from BOLT-LMM, for different values of *K*. In Figure **a** hyperparameter sets 1-8 (all with max depth=2) from Table 4 in the model fitting process are used. In Figure **b** hyperparameter sets 9-16 (max depth=3) are used. Both figures show that the rankings based on the ranking process gives in general better model performance than for the BOLT-LMM ranking. There is also some gain in performance by increasing the hyperparameter max depth from two to three.

**Figure 10:**
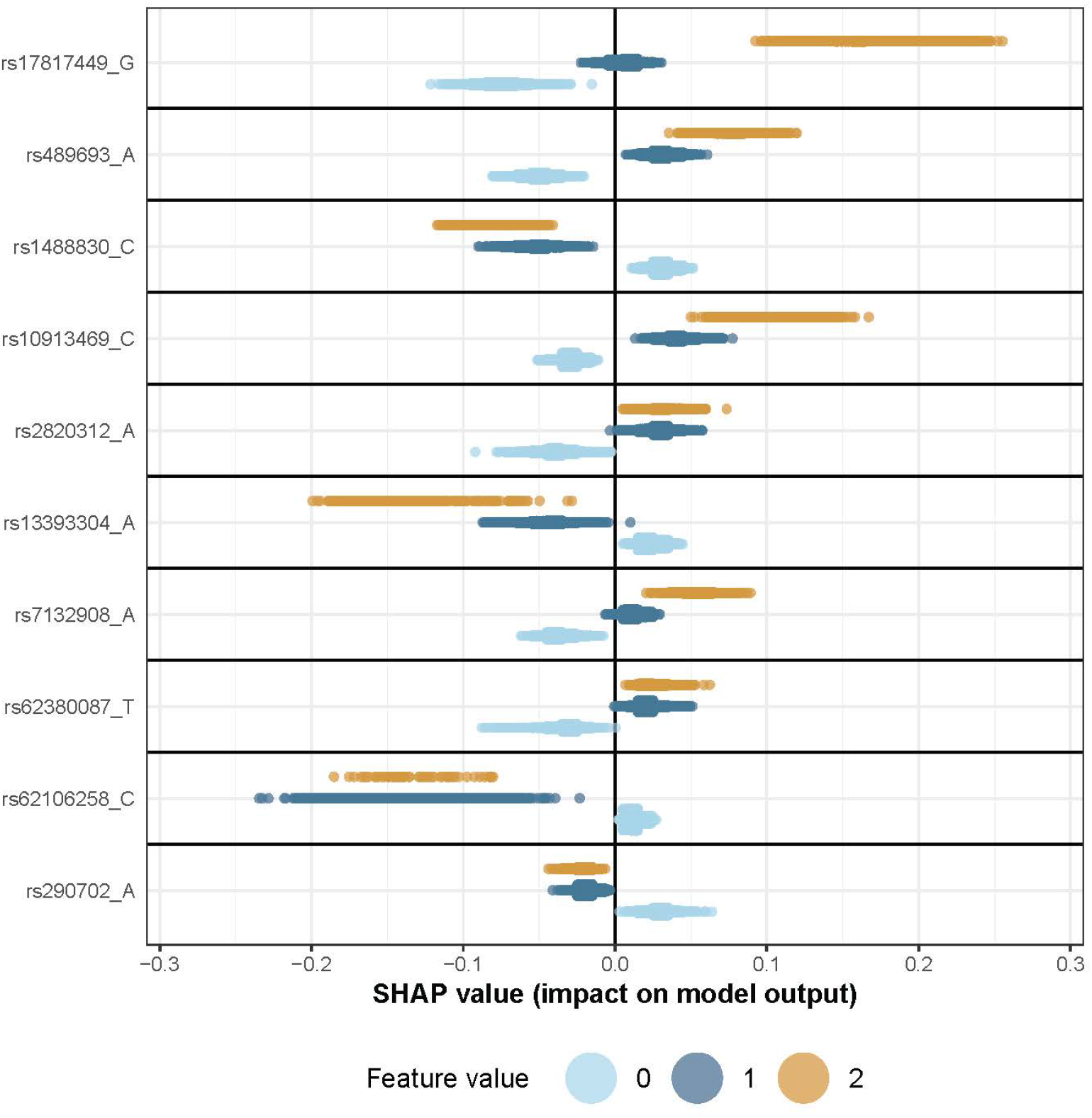
A sina plot visualise the importance of each feature from a fitted model. Here we show the sina plot of the marginal effects for one of the four models described in Section 4.5 when applied to the evaluation data.

**Figure 11:**
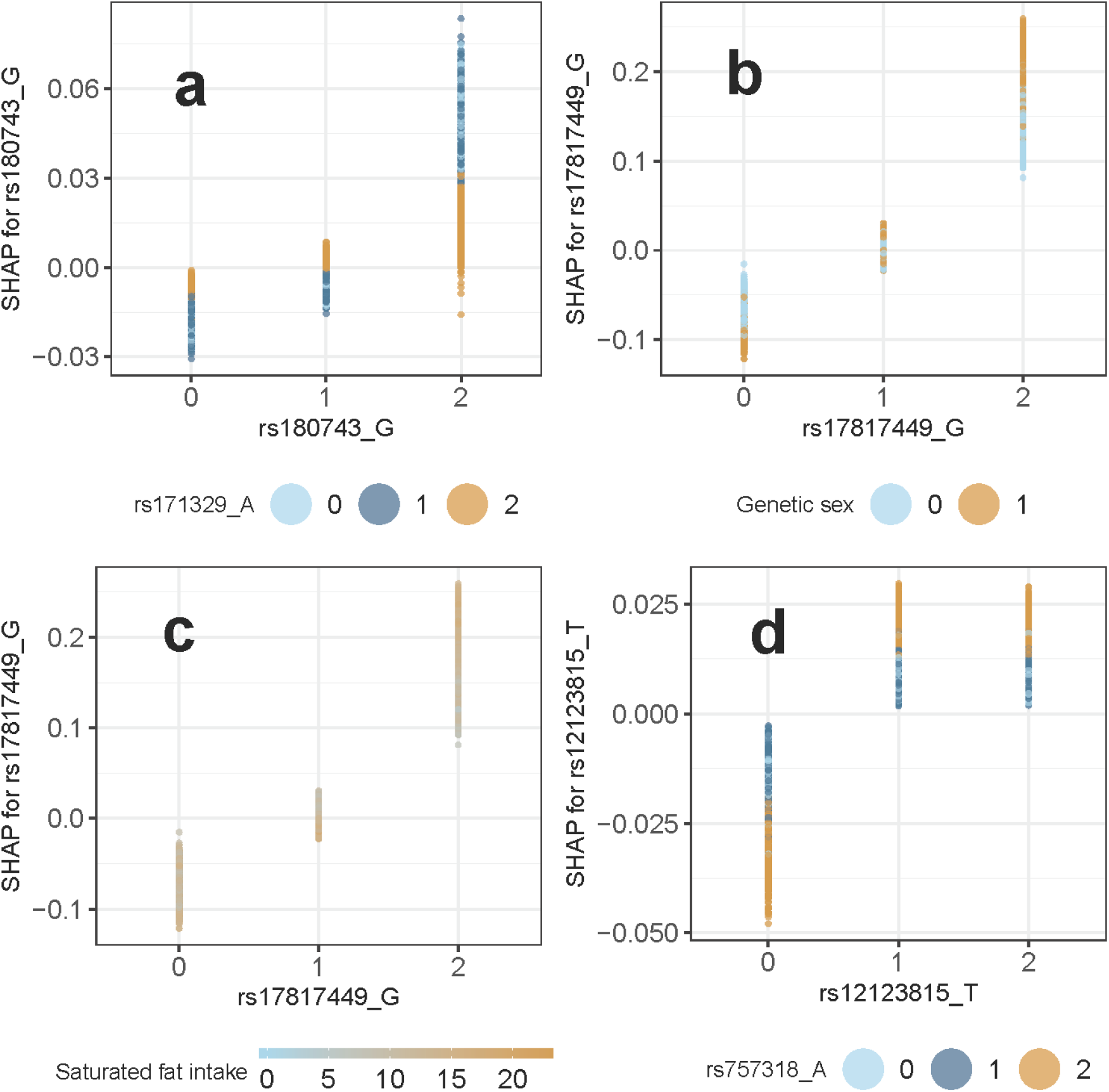
Partial dependence plots for the pairs **a**) rs180743 and rs171329, **b**) rs17817449 and genetic sex, **c**) rs17817449 and saturated fat intake, and **d**) rs12123815 and rs12123815. In all panels we see how the SHAP values (vertical axis) depends on the feature value of the SNP (horizontal axis) and on the value of the second feature (color).

**Figure 12:**
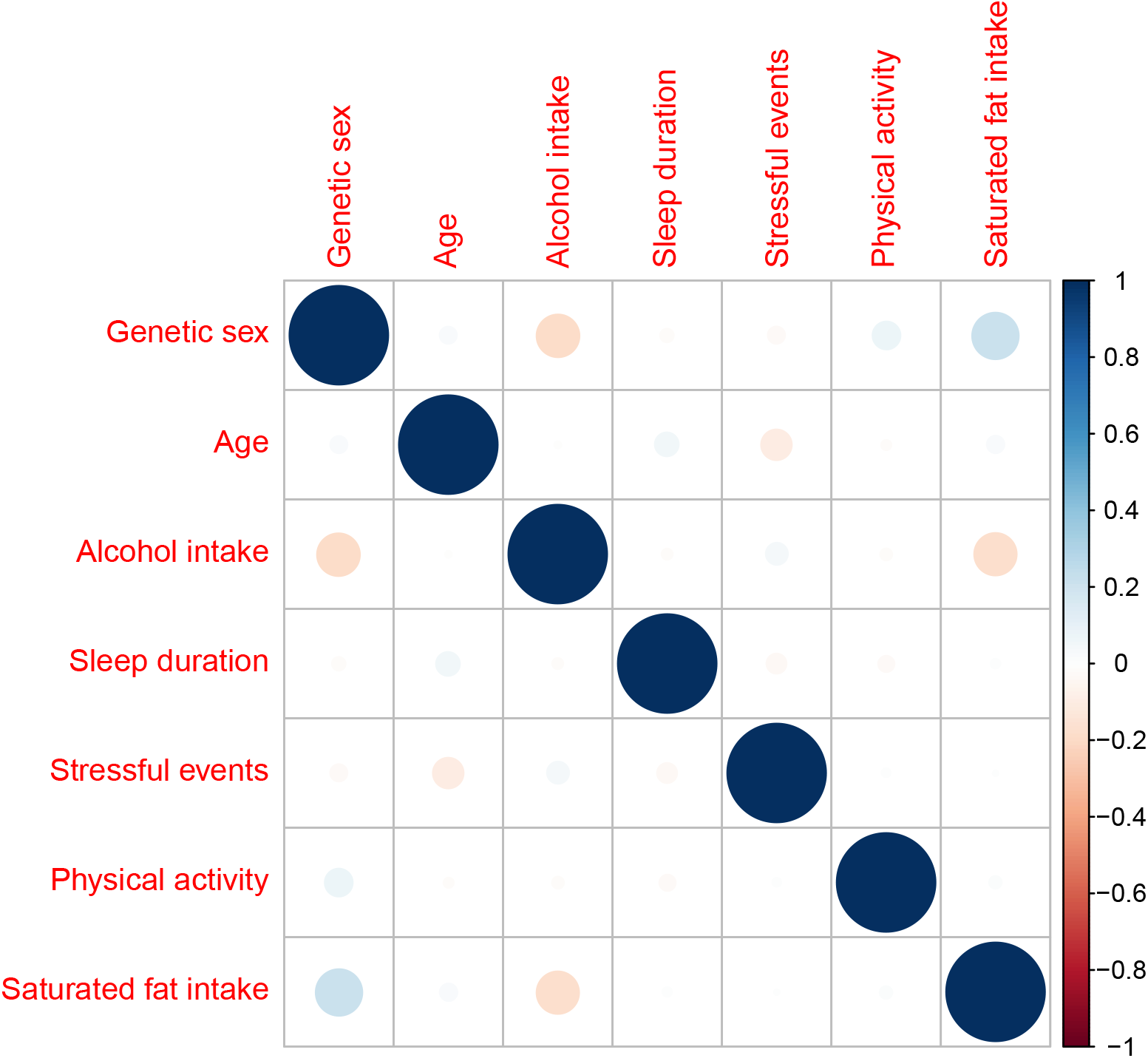
Pearson’s correlation, *r*, between environmental covariates.

The turning point for the BOLT-LMM ranking is *K* = 1000 while for the models based on the ranking process, the turning point is consistently for a larger *K* value. The maximum average PR-AUC-score for the XGBoost models created in cross-validation is in general larger when using the ranking based on our method than the ranking based on BOLT-LMM. From Figure 9, the maximum average performance score is in general better when allowing the regression trees to be of maximum depth three instead of two. Additionally, inclusion of the SNPs provide only a small contribution to the increase in the average prediction performance, where the best models increase the average PR-AUC score from 0.606 when only environmental covariates are included to 0.629 when the top 5000 SNPs are included (blue line, Figure 9**b**). This corresponds to an increase in average classification accuracy from 0.64 to 0.66.

### 4.6 Phase 3: Model explainability

In the model explainability phase we use the evaluation data consisting of 47 015 individuals, that has not been used in Phase 1 and 2. For convenience, we consider the models constructed during cross-validation that performed best on average on the test data during the model fitting process. These are the four models from 4-fold cross-validation trained on the top 5000 ranked features with hyperparameter set 2 visualised as the blue line in Figure 9**b**. We now explore what these four models consider important with respect to their predictions on the evaluation data. This is done as described in Section 3.3 by computing the expected relative contribution for both individual features as well as interactions. Marginal and interaction effects can be visualized with sina plots and partial dependence plots respectively. For the case of marginal effects, Figure 10 shows the sina plot for one of the four models trained on the SNPs with the largest expected relative contributions.

We use Equation (10) together with Equation (12) to compute the average relative interaction contribution (ERIC) for each pair of features based on the evaluation data. The top 10 interactions are given in Table 5.

**Table 5:**
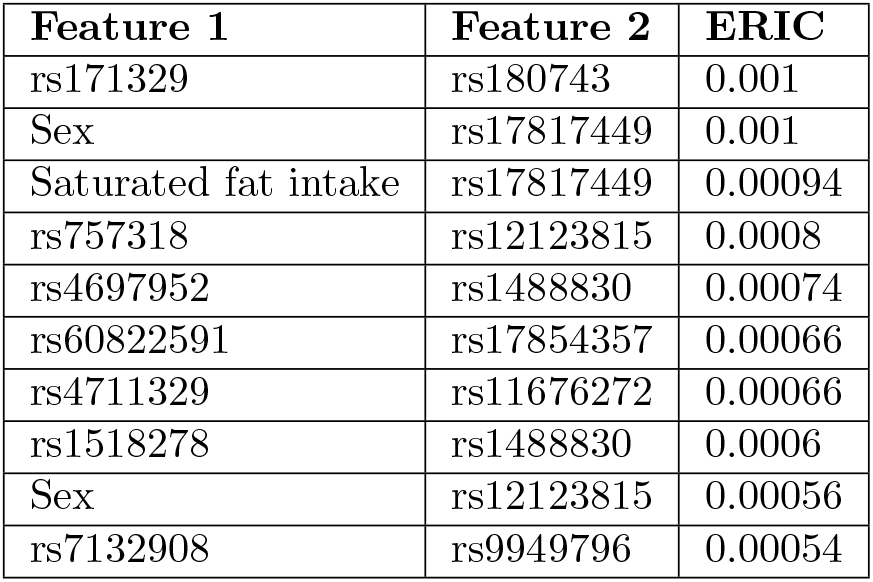
The top 10 interactions based on the expected relative interaction contribution (ERIC) estimated on the evaluation data (Phase 3), with the aim of explaining the best predictive models from Phase 2.

| Feature 1 | Feature 2 | ERIC |
| --- | --- | --- |
| rs171329 | rs180743 | 0.001 |
| Sex | rs17817449 | 0.001 |
| Saturated fat intake | rs17817449 | 0.00094 |
| rs757318 | rs12123815 | 0.0008 |
| rs4697952 | rs1488830 | 0.00074 |
| rs60822591 | rs17854357 | 0.00066 |
| rs4711329 | rs11676272 | 0.00066 |
| rs1518278 | rs1488830 | 0.0006 |
| Sex | rs12123815 | 0.00056 |
| rs7132908 | rs9949796 | 0.00054 |

First of all, we see that the contributions from the interactions, are quite small as no interaction has expected relative interaction contribution more than 0.001. The behaviour of these interactions can be visualized by using partial dependence plots [19, 32]. Figure 11 show the partial dependence plots for the top four interactions from Table 5 when regarding one specific chosen model, out of the four, for each interaction.

We see in Figure 11 examples where the SHAP value of the feature for each individual represented along the x-axis not only depends on its own feature value, but the value of some other feature as well. For instance, in Figure 11**a**, we see that the increased risk of being obese when the genotype value is equal to two for rs180743, is reduced if the genotype value of rs171329 is equal to two as well. We also see in Figure 11**b** that being a male (orange points) is in general more protective against obesity compared to females when the genotype value of rs17817449 is zero. However, males have, in general, a larger risk of being obese than females when the genotype value is two. A positive SHAP value implies a positive contribution to the log-odds prediction, and therefore a contribution making it more likely to be a case (obese).

### 4.7 Interaction models in logistic regression

We compare the interaction rankings from Phase 3 with logistic regression fits on the full UK Biobank dataset and the evaluation data alone. We consider a parametric model, assuming additive effects, for both SNP-SNP and SNP-environment interaction effects for logistic regression, and construct a hypothesis test to infer the presence of interactions. For the test of SNP-SNP interactions between two SNPs *a* and *b*, the null model will be:

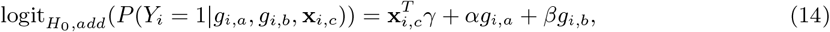

where 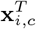 is a vector of features such as intercept, age, environmental features and principal components, while *γ* is the vector of corresponding parameters for each covariate. The parameters *α* and *β* are the marginal effects from SNP *a* and *b* resepectively. The corresponding alternative model will be:

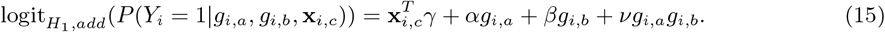

For a SNP-environment interaction we will use the following alternative model:

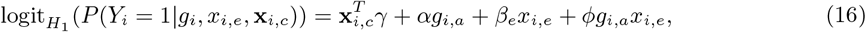

where *β*_*e*_ and *ϕ* are marginal environmental effect and interactions parameters respectively.

For the testing of the interactions we apply the likelihood ratio test (LRT) to test the null hypothesis that *ν* = 0 for SNP-SNP interactions or *ϕ* = 0 for SNP-environment interactions [19, 54]. The LRT assumes independence between the samples, and so we need to make sure the individuals included in the test are not related to any significant degree.

#### 4.7.1 Comparison of Phase 3 results with logistic regression tests

Let the vector **x**_*i,c*_ given in (15) consist of the intercept in addition to the features sex, age and the top four principal components for each individual. The principal components are used to correct for population stratification [16]. The ranking of the pairwise interactions is based on the evaluation data consisting of 47 015 individuals. We fit a logistic regression model based on all unrelated individuals in the evaluation data (39286 individuals), as well as a logistic regression based on all unrelated individuals used in this paper (173468 individuals). Unrelatedness is ensured by using data field 22020 in the UK Biobank Data Showcase [7]. The principal components were calculated using EIGENSOFT (version 6.1.4) SmartPCA [40, 36]. We compute the principal components on the unrelated individuals in the evaluation data and all unrelated individuals separately. PCA plots for both the evaluation data and the full data set can be seen in the Supplementary File. A few individuals have missing values for each test and are removed.

The top four interactions from the SHAP values visualized in Figure 11 are evaluated by applying LRT tests for each interaction. The results are given in Table 6.

**Table 6:** Results from likelihood ratio tests applied on the top four ranked interactions found from the model explainability process based on the evaluation data.

| Data set | Interaction | $p$-value LRT |
| --- | --- | --- |
| Evaluation data | rs171329 and rs180743 | 0.85 |
| All individuals | rs171329 and rs180743 | 0.024 |
| Evaluation data | rs17817449 and genetic sex | 0.77 |
| All individuals | rs17817449 and genetic sex | 4.09e-05 |
| Evaluation data | rs17817449 and saturated fat intake | 0.44 |
| All individuals | rs17817449 and saturated fat intake | 0.0017 |
| Evaluation data | rs757318 and rs12123815 | 0.25 |
| All individuals | rs757318 and rs12123815 | 0.71 |

It is clear that the sample size is the dominating factor for the computed *p*-values. All *p*-values based on the evaluation data, the same data that is used to rank the interactions, are non-significant. As expected, the *p*-values are in general smaller when considering all individuals, yet none of them would be declared significant in the case of any reasonable multiple testing procedure [17].

The smallest *p*-value is achieved for the interaction between the SNP rs17817449 and genetic sex when including all individuals. In the Supplementary File, we apply likelihood ratio tests based on logistic models with less stricter additive assumptions. However, less stricter additive assumptions do not provide smaller p-values to any significant degree.

## 5 Discussion

We have proposed how tree ensemble models, such as implemented in XGBoost, can be combined with SHAP values to explain the importance of individual features (SNP or environmental factor) for a specific trait as well as for identification of interactions between features. The method has been illustrated on an example from the UK Biobank. We have shown that through several independent cross-validations on XGBoost models using subsets of SNPs spread along the genome, one is able to find a reasonable ranking of individual features similar to what is found in previous GWAS of obesity [25].

### 5.1 Ranking and interactions

When comparing the SNP ranking from the SHAP values to the BOLT-LMM approach with ranking through *p*-values, Figure 9 suggests that the ranking process has the potential to outperform BOLT-LMM in predictive power, due to non-linear effects not detected through BOLT-LMM.

The SHAP values can also be used to identify interactions. Comparing the top ranked interactions with classical logistic regression including interaction parameters, we see that none of the corresponding statistical tests provide convincing *p*-values. Assuming the ranking of interactions via SHAP values is reliable, we see from Table 5 that the interaction effects are small, even though we can see clear patterns in Figure 11. The non-significant *p*-values can be a direct result of small effect sizes together with a small sample size. Additionally, potential parametric logistic regression models needed to capture these interactions require a larger number of *degrees of freedom* which can reduce statistical power [54]. However, there is a need to develop tests that can infer the trustworthiness of the results found from SHAP values in a similar fashion as through *p*-values in classical statistical theory.

### 5.2 Data split

In this paper, data is split in three subsets used for ranking, model fitting and model explanation respectively. This procedure requires a large amount of data, but the purpose was to evaluate the credibility and potential of using tree ensemble models together with SHAP values. For smaller data samples, an alternative procedure is to rank interactions directly during the ranking process by computing the expected relative interactions contributions explained in Section 3.3. However, the ranking process consists of many models with low predictive power, which makes it more difficult to explore the true relationships compared to the models constructed in the model fitting process.

### 5.3 Limitations and improvements

The choice of number of SNPs *S*, individuals *G*, folds *F* and *r*^2^-threshold in each cross-validation in the ranking process are all important with respect to performance, and should be considered as hyperparameters. The number of SNPs *S* must be large enough to represent important regions in the genome, but not so large that it introduces noise to the model. The number of individuals in each cross-validation, *G*, should be as large as possible as it increases the power to detect small as well as nonlinear effects. However, that may lead to computational challenges. The number of folds in the cross-validations, *F*, should neither be too small nor too large as we want to train the model on as many different subsets of the population as possible in order to find the most general effects, but at the same time the validation dataset must be large enough to be sufficiently representative.

The mutual independence assumption when computing the SHAP values is a significant restriction, and a mutual *r*^2^ below any threshold between features will by no means ascertain mutual independence as *r*^2^ measures linear dependency. Correlation measures that can also account for non-linear dependencies in a high-dimensional setting could provide more trustworthy results.

Another issue is the measure of the expected relative contribution for both marginal effects and interaction effects given in Equations (9) and (12). These measures depend on how many individuals are evaluated to compute the SHAP values in each model given by *G*_*p*_. In a heterogeneous population, *G*_*p*_ should most likely be larger than the sizes used in this paper. Metrics for how representative a subset is to all data would be beneficial to decide the optimal size.

It is also important to investigate the extent to which cross-validation reduces the negative effects of population stratification and cryptic relatedness and then incorporate improvements that can take into account these effects. For instance, to address population stratification it would be interesting to investigate how principal components as features could be used in the tree ensemble models.

### 5.4 Hyperparameter optimization

We have seen that the hyperparameters for XGBoost are important. Unfortunately, the computation time for each set of hyperparameters is protracted, and consequently systematic hyperparameter optimization is not feasible. However, from the choice of hyperparameter sets in this paper, the hyperparameters col-sample bytree, subsample and colsample bylevel should be high (0.8-0.9) while the learning rate *η* should be low (0.05-0.1), but not too low. Another important hyperparameter, the regularization parameter, *λ* should be investigated more extensively.

### 5.5 Predictive performance and obesity

Even with strong predictors such as physical activity, intake of saturated fat, alcohol use, stressful events, sleep duration, age and sex in addition to genome-wide genetic data, we are not capable of constructing a model with more than 66% classification accuracy, and the genetic data only provide a small portion of the predictive performance. The usefulness lies in the fact that tree ensemble models such as XGBoost, unlike a linear mixed model, are capable of identifying non-linear effects. However, in the example of obesity, these non-linearities (in the log-odds scale) seem not to be of great importance. If the prediction performance of the model is considered satisfactory, this can be an important diagnostic tool in the future.

## 6 Conclusion

We have seen that our proposed tree ensemble- and SHAP-based method gives us the possibility of detecting both gene-gene and gene-environment interactions that are not detected using test for interactions in logistic regression. Our proposed method can be applied to high-dimensional genetic data in large-scale biobanks. There is however a need to develop methods for assessing the variability of SHAP interaction values, and to develop a hypothesis test for SHAP interaction values.

## Data Availability

The research has been conducted using the UK Biobank Resource under Application Number 32285.

## Acknowledgements

This research was supported by the Norwegian Research Council grant 272402 (PhD Scholarships at SINTEF) as well the funding for research stays abroad for doctoral and postdoctoral fellows financed by the Norwegian Research Council. The research has been conducted using the UK Biobank Resource under Application Number 32285. We thank the Yale Center for Research Computing for guidance and use of the research computing infrastructure. We thank the The Gemini Center for Sepsis Research for establishing cooperation with Yale School of Public Health.

## Code availability

The source code supporting this paper can be found online at https://github.com/palVJ/GWASwithTreeSHAP/.

## A Quality assessment of UK Biobank Genetic Data

Analyses were limited to autosomal variants covered by both genotype arrays used over the course of the study and that passed the batch-level quality control. SNPs were included if the call rate was above 99%, the Hardy-Weinberg equilibrium *p*-value was less than 5 · 10^*−*8^, and the minor allele frequency was larger than 1%. 529,024 SNPs passed these filters.

Individuals were removed if the genetic and reported sex did not match and if the sex chromosomes were not XX or XY. Outliers in heterozygosity and missing rates were removed. The analyses were limited to those identified as Caucasian through the UK Biobank’s PCA analysis (field 22006). All individuals had an individual call rate larger than 99%. 366,752 individuals passed these filters.

## B Details of environmental covariates from UK Biobank

A sample set of personal and environmental characteristics were included in the model as covariates to demonstrate sample use of the method. All descriptions are from the UK Biobank Showcase, and no outliers were removed. Individuals that answered “prefer not to answer” or “do now know” to any given question were treated as missing values. All features are taken from the baseline assessment, the same point in time when the BMI phenotype was measured. The following environmental and personal features collected at baseline were evaluated:

| Description | Data field |
| --- | --- |
| Age when attended assessment centre | 21003 |
| Genetic sex | 22001 |
| Number of days/week walked 10+ minutes | 864 |
| Minutes spent walking per day | 874 |
| Number of days/week of moderate physical activity 10+ minutes | 884 |
| Duration of moderate activity per day | 894 |
| Number of days/week of vigorous physical activity 10+ minutes | 904 |
| Duration of vigorous activity per day | 914 |
| Alcohol intake frequency | 1558 |
| Sleep duration | 1160 |
| Processed meat intake | 1349 |
| Beef intake | 1369 |
| Lamb/mutton intake | 1379 |
| Pork intake | 1389 |
| Cheese intake | 1408 |
| Milk type used | 1418 |
| Illness, injury, bereavement, stress in last 2 years | 6145 |

### B.1 Age when attended assessment centre

Age at the initial assessment visit (2006-2010) during which participants were recruited and provided consent.

### B.2 Genetic sex

Sex as determined from genotyping analysis.

### B.3 Physical activity

To measure the degree of physical activity, the duration of walking, moderate activity and vigorous activity per day were added with equal weight. The duration of any given activity per day is set to zero if an individual spent no days during the week with more than 10 minutes of that activity.

### B.4 Alcohol intake

Participants were asked how frequently they consumed alcohol, with potential responses never, only on special occasions, one to three times a month, one to three times a week, three or four times a week, or daily or nearly daily.

### B.5 Sleep duration

Participants were asked to report how many hours of sleep they got in a 24 hour period.

### B.6 Saturated fat intake

Participants were asked how frequently they consumed each food item, from never to daily. Frequency of beef, lamb, mutton, pork, cheese and milk intake per week was added with equal weight.

### B.7 Stressful events

We treated this as a binary variable, such that those that have not experienced any of the categories listed in the “Illness, injury, bereavement, stress in last 2 years” variable during the past two years are represented by the value zero, and the rest were set to one.

### B.8 Treatment of categorical features and correlation plot

XGBoost does not automatically take into account categorical features. Sex, alcohol consumption and sleep duration can be considered categorical features, but as sex is a binary feature, while alcohol consumption and sleep duration are ordinal features, a split between two categories for these features in a regression tree is meaningful, and therefore the features can be treated as they are. The correlation of the final seven environmental covariates were investigated further by computing the Pearson’s correlation between all pairs of covariates by excluding missing values. No pairs of covariates showed Pearson’s correlation *r* larger than 0.2, and we therefore treat these covariates as if they were independent of each other when computing the SHAP values. Correlations between environmental covariates and SNPs are also surprisingly not very small. Even though there exist dependence between SNPs and environmental covariates, the effects are so small that we also in this case regard them to be independent to each other when computing the SHAP values.

